# Gene-Gene and Gene-Environment Interactions Improve Genetic Risk Predictions for Complex Diseases

**DOI:** 10.64898/2026.08.12.26360291

**Authors:** Keri Multerer, Yosuke Tanigawa, Manolis Kellis, Lisa Woods, Binh Nguyen, Paul H. Atkinson, Andrew B. Munkacsi

## Abstract

Polygenic risk scores (PRS) assume additive SNP effects, yet genetic risk also arises from interactions between loci and environmental factors that contribute to broad-sense heritability. We developed an extended PRS (ePRS) framework for type 2 diabetes (T2D) that incorporates locus-by-locus non-additive effects beyond those captured by additive single-locus PRS or linkage disequilibrium (LD) tagging. These were modelled as interaction-informed cumulative burden (G+G; summed allele counts), epistasis-screened (G*×*G; allele count products), and gene-environment effects derived from cardiometabolic variables in electronic health records. Across 235,000 UK Biobank participants, five complementary ePRS models captured largely non-overlapping high-risk individuals, suggesting that a key to individual risk predictions comprises the inclusion of multiple interaction-driven biological components rather than a single signal. A composite score was predictive over traditional additive PRS and predictive of T2D among individuals whose clinical symptoms fall within normal ranges.

## 1 Introduction

Polygenic risk scores (PRS) estimate genetic risk by aggregating additive effects from genome-wide association studies (GWAS) [1–6], despite widespread recognition that complex disease risk reflects broad-sense heritability (H^2^) and includes gene-gene and gene-environment interactions in addition to additive effects [7–11]. Although numerous methods have sought to model higher-order genetic effects [12–18], most either do not explicitly retain interaction terms or sacrifice explainability for predictive performance [19]. Consequently, the contribution of interaction-aware features to clinically useful PRS remains unclear [9, 20, 21].

We developed an interpretable machine-learning framework that incorporates complementary multi-locus feature encodings into PRS while retaining explicit loci and environmental variable contributions. A Cornerstone of our framework is the systematic comparison of G+G and G*×*G allele encodings derived from identical SNP pairs preselected using PLINK’s [22] computationally efficient epistasis screening. The first, *interaction-informed multilocus additive burden* (G+G), sums allele counts across paired loci to capture cumulative allelic burden that may be undetected by traditional marginal single-SNP analyses. The second, *epistasis-screened* (G*×*G), encodes allele-count effect products to model departures from additive independence. These measurements were further combined with cardiometabolic variables collected at the UK Biobank initial assessment to generate corresponding gene-environment interaction models (G+G*×*E and G*×*G*×*E). Unlike approaches that model higher-order effects through complex algorithms without identifying specific interactions or their combined effect sizes, our framework preserves explicit interaction features, enabling direct biological interpretation [23–26].

We then evaluated whether these encodings capture complementary components of polygenic risk that traditional additive PRS alone would not account for and whether incorporating environmental interactions (cardiometabolic clinical measures) improves prediction beyond genetic information alone. To answer this, we applied our extended PRS (ePRS) framework to type 2 diabetes (T2D) and coeliac disease (CD), two diseases with contrasting genetic architectures. Thus, T2D is highly polygenic, with risk distributed across hundreds of variants influencing insulin secretion, insulin resistance and metabolic regulation, whereas CD is dominated by large human leukocyte antigen (HLA) effects, a comparatively defined genetic architecture. Comparing these diseases in this way enabled us to assess how multi-locus feature encoding performs across such distinct genetic architectures.

We generated five statistically distinct ePRS models (PRS*_G_*, PRS*_GxG_*, PRS*_G_*_+*G*_, PRS*_G_*_+*GxE*_ and PRS*_GxGxE_*), that included interaction-aware feature encodings thus identifying largely non-overlapping high-risk individuals for future T2D: 4,187 (78%) of 5,392 UK Biobank cases in the validation set were identified, with 2,200 (53%) cases exclusive to a single model. A composite score (ePRS_Comp_) combining interaction models improved discrimination over traditional additive PRS and were predictively informative of T2D among individuals within clinical normal measures, however its incremental value beyond clinical assessment was limited as those measures identified most T2D cases. In coeliac disease, where large additive HLA effects dominate, the traditional additive score carried comparatively more of the PRS value, indicating that the benefit of ePRS modelling must also account for genetic architecture.

## 2 Results

### 2.1 Characterizing ePRS models with G+G, G*×*G, G+G*×*E, and G*×*G*×*E feature encoding

We developed an extended PRS (ePRS) framework to investigate non-linear genetic and gene–environment effects in type 2 diabetes (T2D), a highly polygenic disease with substantial environmental contributions. We also validated the framework performance in coeliac disease, which has a distinct genetic architecture. We analysed *∼*624,000 directly genotyped variants (G) (Methods 4.1), *∼*608,388 significant G*×*G pairs (*p <* 5 *×* 10*^−^*^6^), 181 imputed HLA allelotypes replacing SNPs within the MHC region[27], and 72 cardiometabolic clinical measures with *<* 5% missingness (Extended Table S12). Our pipeline incorporated novel feature reduction and gene-environment discovery methods (Methods 4.5.3). The performance characteristics of our four new ePRS models are shown in Table 2.

**Table 1:** Analysis of pairwise model redundancy for type 2 diabetes and coeliac disease. Redundant pairs (highlighted in red) failed *≥*2 of 3 distinctness criteria. *κ* = Cohen’s Kappa [35]; Jaccard = Jaccard overlap index [36]; DeLong *p* = *p*-value from DeLong AUC comparison [37]; ΔAUC = absolute AUC difference; Criteria = number of distinctness criteria met. *κ* and Jaccard are computed among cases at the top-20% high-risk threshold; DeLong *p* and ΔAUC are threshold-free. Criterion 3 requires both DeLong *p <* 0.05 and |ΔAUC*| >* 0.05.

| Model comparisons | $\kappa$ | Jaccard | DeLong $p$ | $\Delta\text{AUC}$ | Criteria | Decision |
| --- | --- | --- | --- | --- | --- | --- |
| **Type 2 Diabetes** |  |  |  |  |  |  |
| $\text{PRS}_G$ vs $\text{PRS}_{GxG}$ | 0.01 | 0.15 | $<0.00$ | 0.07 | 3/3 | Distinct |
| $\text{PRS}_G$ vs $\text{PRS}_{GxGxE}$ | -0.01 | 0.17 | 0.013 | 0.01 | 2/3 | Distinct |
| $\text{PRS}_G$ vs $\text{PRS}_{G+G}$ | 0.30 | 0.34 | 0.30 | 0.00 | 2/3 | Distinct |
| $\text{PRS}_G$ vs $\text{PRS}_{G+(G+G)}$ | 0.73 | 0.65 | 0.21 | 0.01 | 0/3 | Redundant |
| $\text{PRS}_G$ vs $\text{PRS}_{G+(GxG)}$ | 0.51 | 0.45 | $2.6 \times 10^{-11}$ | 0.02 | 1/3 | Redundant |
| $\text{PRS}_G$ vs $\text{PRS}_{G+GxE}$ | 0.01 | 0.18 | $<0.00$ | 0.02 | 2/3 | Distinct |
| $\text{PRS}_{GxGxE}$ vs $\text{PRS}_{G+GxE}$ | -0.03 | 0.16 | $<0.00$ | 0.03 | 2/3 | Distinct |
| $\text{PRS}_{GxG}$ vs $\text{PRS}_{G+G}$ | 0.02 | 0.15 | $<0.00$ | 0.07 | 3/3 | Distinct |
| **Coeliac Disease** |  |  |  |  |  |  |
| $\text{PRS}_G$ vs $\text{PRS}_{GxG}$ | 0.15 | 0.32 | $<0.00$ | 0.16 | 3/3 | Distinct |
| $\text{PRS}_G$ vs $\text{PRS}_{GxGxE}$ | 0.23 | 0.45 | $<0.00$ | 0.12 | 3/3 | Distinct |
| $\text{PRS}_G$ vs $\text{PRS}_{G+GxE}$ | 0.24 | 0.45 | $<0.00$ | 0.12 | 3/3 | Distinct |
| $\text{PRS}_G$ vs $\text{PRS}_{G+G}$ | 0.70 | 0.78 | 0.10 | 0.01 | 0/3 | Redundant |
| $\text{PRS}_G$ vs $\text{PRS}_{G+(G+G)}$ | 0.78 | 0.83 | 0.10 | 0.01 | 0/3 | Redundant |
| $\text{PRS}_{GxG}$ vs $\text{PRS}_{G+(GxG)}$ | 0.94 | 0.89 | $4.5 \times 10^{-9}$ | 0.00 | 0/3 | Redundant |
| $\text{PRS}_{GxGxE}$ vs $\text{PRS}_{G+GxE}$ | 0.40 | 0.50 | 0.74 | 0.00 | 0/3 | Redundant |

**Table 2:** Performance metrics for statistically distinct ePRS models in the validation and holdout sets for T2D and CD.

| Metric | PRS<sub>G</sub><br>(additive) | PRS<sub>G+G</sub><br>(G+G burden) | PRS<sub>GxG</sub><br>(G×G epistasis) | PRS<sub>G+GxE</sub><br>(G+G×E) | PRS<sub>GxGxE</sub><br>(G×G×E) | ePRS<sub>Comp</sub><br>(fitted combination) |
| --- | --- | --- | --- | --- | --- | --- |
| **Type 2 Diabetes** |  |  |  |  |  |  |
| AUC, PRS alone (95 % CI)<sup>††</sup> | 0.59 (0.58–0.60) | 0.59 (0.58–0.60) | 0.52 (0.51–0.53) | 0.57 (0.56–0.58) | 0.60 (0.60–0.61) | 0.66 (0.65–0.67)** |
| AUC, PRS alone, holdout (95 % CI)<sup>#</sup> | 0.58 (0.57–0.59) | 0.59 (0.58–0.60) | 0.51 (0.50–0.52) | 0.56 (0.54–0.57) | 0.60 (0.59–0.61) | 0.65 (0.64–0.66) |
| Nagelkerke $R^2$ (95 % CI)<sup>‡</sup> | 0.017 (0.014–0.020) | 0.017 (0.014–0.020) | 0.001 (0.000–0.001)<sup>§</sup> | 0.015 (0.012–0.017) | 0.006 (0.004–0.008) | 0.037 (0.031–0.044)<sup>##</sup> |
| Precision (recall) | 0.12 (0.29) | 0.12 (0.29) | 0.09 (0.22) | 0.11 (0.29) | 0.12 (0.29) | 0.14 (0.36) |
| Cases in top 20% | 1,569 | 1,555 | 1,166 | 1,539 | 1,574 | 965 |
| Cases not identified by PRS<sub>G</sub><sup>†</sup> | — | +765 (49%) | +818 (52%)<sup>§</sup> | +1,076 (69%) | +1,125 (72%) | +466 (61%) |
| Exclusive cases | 297 | 301 | 341 | 516 | 745 | — |
| N features | 809 | 527 | 1,200 | 27 | 17 | — |
| N HLA loci | 85 | 102 | 112 | 3<sup>¶</sup> | 1<sup>¶</sup> | — |
| **Coeliac Disease** |  |  |  |  |  |  |
| AUC, PRS alone (95 % CI)<sup>††</sup> | 0.75 (0.74–0.76) | *excl.\** | 0.58 (0.56–0.61) | 0.63 (0.61–0.65) | *excl.\** | 0.75 (0.73–0.77)** |
| AUC, PRS alone, holdout (95 % CI)<sup>#</sup> | 0.74 (0.71–0.76) | *excl.\** | 0.59 (0.56–0.62) | 0.65 (0.61–0.68) | *excl.\** | 0.75 (0.72–0.78) |
| Nagelkerke $R^2$ (95 % CI)<sup>‡</sup> | 0.092 (0.077–0.107) | — | 0.008 (0.005–0.013) | 0.019 (0.011–0.029) | — | 0.036 (0.020–0.059)<sup>##</sup> |
| Precision (recall) | 0.03 (0.59) | — | 0.02 (0.29) | 0.02 (0.43) | — | 0.03 (0.60) |
| Cases in top 20% | 441 | — | 219 | 325 | — | 224 |
| Cases not identified by PRS<sub>G</sub><sup>†</sup> | — | — | +60 (14%) | +88 (20%) | — | +20 (10%) |
| Exclusive cases | 132 | — | 46 | 74 | — | — |
| N features | 178 | — | 2,998 | 102 | — | — |
| N HLA loci | 59 | — | 49 | 38<sup>¶</sup> (13 unique) | — | — |

<sup>†</sup>Cases in this model’s top 20% not in the PRS<sub>G</sub> top 20%, with the percentage of PRS<sub>G</sub>-identified cases in parentheses.
<sup>‡</sup>Incremental Nagelkerke $R^2$ above a covariate-only model; 95% CI from 2,000 bootstrap resamples where available.
<sup>§</sup>Despite lower AUC and $R^2$ , PRS<sub>GxG</sub> captures 818 T2D cases not found with PRS<sub>G</sub> top 20%.
<sup>||</sup>Cases uniquely identified by this model.
\* excluded as statistically redundant (Table 1).
<sup>¶</sup>HLA included in G×G×E and G+G×E features and not modelled separately.
<sup>††</sup>AUC is for the unadjusted ePRS (T2D $n = 67,402$ ; $N_{\text{cases}} = 5,392$ ; CD $n = 67,404$ ; $N_{\text{cases}} = 748$ ).
\*\*ePRS<sub>Comp</sub> weights were fitted in the validation cohort, with AUC as 5-fold cross-validated within that cohort while the holdout value is fully out-of-sample; validation-holdout differences are 0.009 in T2D and 0.005 in CD.

From our UK Biobank dataset, we used 70% of participants (*n* = 235,956) for model training and feature reduction, 20% (*n* = 67,402) for validation, and 10% (*n* = 33,712) as an independent holdout (Methods 4.2) of unrelated white British participants[28]. The validation set was used to calculate ePRS scores and select models, while the untouched holdout was used for final performance evaluation and construction of ePRS_Comp_. Feature reduction identified 915 G, 1,497 G+G, and 13,767 G*×*G features for subsequent G*×*E discovery (Methods 4.5.3). This yielded 304 G+G*×*E interactions (69 unique environmental variables) and 1,462 G*×*G*×*E interactions (30 unique environmental variables) (Extended Table S2) used in association analysis. Equivalent results for CD are reported in Table 2. To preserve interaction-specific signals, linkage disequilibrium (LD) pruning was performed after feature selection rather than before (Methods 4.5.2), followed by a secondary departure-from-additivity test to assess whether retained G*×*E terms reflected interaction rather than clinical measure main effects (Methods 4.5.5).

A central feature of our framework was the comparison of two alternative encodings derived from the same significant SNP pairs. G+G encoded cumulative allelic effects by summing allele counts across each SNP pair, whereas epistasis-screened G*×*G encoded by multiplying allele counts to capture departures from additive independence. Following feature reduction, separate feature matrices were generated for each of our models enabling direct comparison of additive allelic effects burden and interaction-based features within the ePRS pipeline.

Seven L1-penalised logistic regression models spanning five feature classes (G, G+G, G*×*G, G+G*×*E and G*×*G*×*E) were trained. Redundancy filtering retained five statistically distinct T2D models (PRS*_G_*, PRS*_G_*_+*G*_, PRS*_GxG_*, PRS*_G_*_+*GxE*_, PRS*_GxGxE_*), representing 2,399 unique single- and interaction-derived loci (Table 1; Extended Tables S2 and S14). The combined models (PRS*_G_*_+(*G*+*G*)_ and PRS*_G_*_+(*GxG*)_) were redundant and excluded from further analysis. Notably, PRS*_G_* remained non-redundant when evaluated separately with either PRS*_G_*_+*G*_ or PRS*_GxG_* but became redundant when combined (PRS*_G_*_+(*G*+*G*)_ and PRS*_G_*_+(*GxG*)_) indicating that single-locus effects provide information distinct from either multi-locus encoding alone.

#### Additional departure from additive analysis

Because the final ePRS models encode each SNP pair as a single feature without separately including the individual SNP effects, we assessed departure from additivity using models that included both loci and their product (Methods 4.5.5). This analysis was restricted to features with non-zero *β*-coefficients in the final ePRS models, thereby testing interactions after accounting for individual locus effects. These analyses inform how ePRS features are described, rather than whether they are retained. Model retention for downstream analyses was based on held-out performance and statistical distinctness between models (Table 1), with coefficients reflecting the predictive contribution of each encoded feature regardless of its underlying source.

Most testable SNP pairs underlying the G+G and G*×*G features showed evidence of departure from additivity. To estimate interactions reliably, we required at least 2,000 individuals carrying both variants. This excluded 187 of 1,088 PRS*_GxG_* pairs (17%) and 19 of 425 PRS*_G_*_+*G*_ pairs (4%) in T2D, compared with 1,000 of 2,949 (34%) and 18 of 229 (8%) in CD, where the constituent variants are rarer. Among the remaining pairs in T2D, 626 of 901 G*×*G pairs (70%) and 233 of 380 G+G pairs (61%) showed significant departure from additivity. In CD, these proportions were higher, with 1,762 of 1,949 G*×*G pairs (91%) and 153 of 211 G+G pairs (73%) showing significant departure. These proportions should not be directly compared between diseases because the higher exclusion rate in CD preferentially removes rarer pairs, leaving a subset with greater power to detect interactions. Per-pair interaction coefficients, standard errors and *q*-values are given in Extended Table S16.

The environmental interaction features showed a different pattern (Extended Table S17). Of the 44 retained T2D environmental interaction features, 41 were testable (all 27 G+G*×*E and 14 of the 17 G*×*G*×*E; the remaining three fell below the double-carrier threshold). None showed statistically significant departure from additivity after correction, although more showed nominal evidence than expected by chance. In CD, 10 of 102 testable G+G*×*E features were significant. Notably, the recovered alleles spanned the established coeliac risk haplotypes, including both chains of the DQ2.5 heterodimer responsible for presenting deamidated gluten peptides and a precursor to the autoimmune response [29] (Extended Table S17). The direction of these associations indicated a greater anthropometric (body measurements) deficit among risk-allele carriers, consistent with established disease rather than evidence of a predictive interaction.

We therefore describe the G*×*E and G*×*G*×*E features as interaction encodings rather than established statistical interactions. ePRS coefficients represent the total predictive effects of the encoded features, rather than partial interaction effects estimated in the confirmatory analyses. For G+G, the encoding is additive in form: the initial PLINK analysis determines which loci are combined, but their allele counts are then simply summed. The 233 of 380 testable pairs that departed from additivity therefore support the epistasis-based selection of loci to combine, while the resulting G+G feature remains additive. Thus, PRS*_G_*_+*G*_ can capture information beyond PRS*_G_* by combining loci selected for evidence of interaction, while retaining an additive allele-count encoding.

The top 20% of each ePRS distribution defined the high-risk group for downstream analyses. A fully independent holdout cohort (*n* = 33,712), excluded from all model development, was used to identify features uniquely predictive of high-risk individuals within each retained ePRS model and to derive the composite score, ePRS*_Comp_*(Section 2.6), which integrates complementary risk captured across the five models. The complete workflow is summarised in Figure 1 which shows additional (to traditional PRS) high risk T2D cases identified in our ePRS analysis .

**Figure 1:**
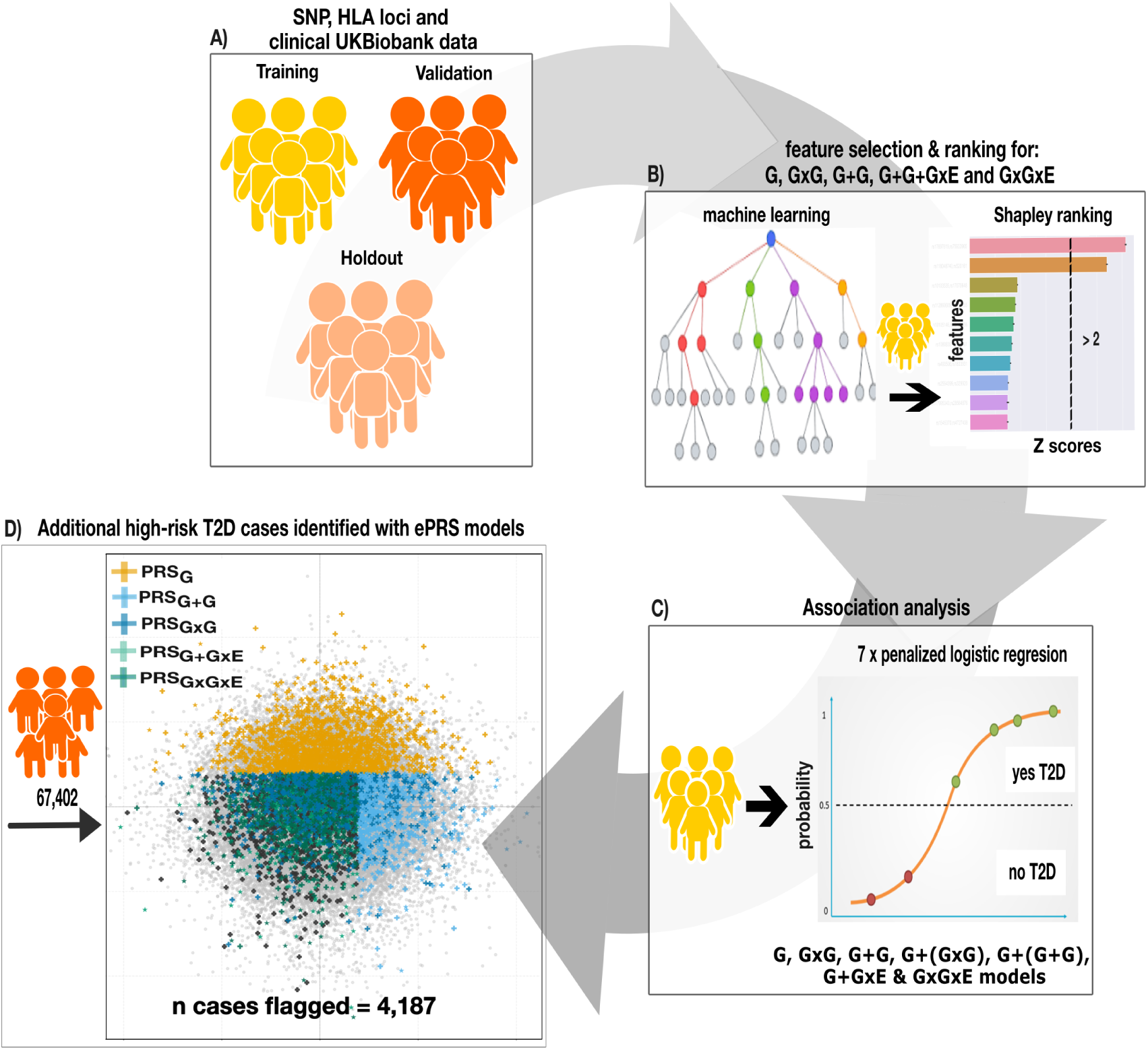
Overall T2D workflow for ePRS with G+G and G*×*G feature encoding. This ePRS pipeline was developed using UK Biobank participants. From top clockwise to bottom: A) the dataset was split into training (70%), validation (20%), and holdout (10%). B) Feature selection was performed separately for G, encoding strategies G+G and G*×*G, and their inclusion in gene-environment analysis, incorporating gradient-boosted classifiers followed by Shapley value ranking to reduce dataset sizes. C) Reduced G and 2 multi-locus feature sets were further combined with cardiometabolic environmental variables to yield G+G*×*E and G*×*G*×*E used to train seven L1-regularised models. D) Non-zero *β* coefficients were used to calculate ePRS with at-risk population in the 20% for each model: PRS*_G_*, PRS*_GxG_*, PRS*_G_*_+*G*_, PRS*_G_*_+*GxE*_ and PRS*_GxGxE_* in validation and holdout data. Five distinct T2D ePRS models reveal complementary risk cohorts. Cases are assigned hierarchically and coloured by identifying model to avoid double counting or black for cases missed with all models. Controls are light-grey circles

To benchmark our approach, we compared the PRS*_G_* model with that of Tanigawa *et al.*[30] using the same UK Biobank training cohort, T2D phenotype definition, and comparable L1-penalised regression framework. In our approach, despite using 3.8-fold fewer variants (809 versus 3,495), PRS*_G_* achieved competitive predictive performance (AUC = 0.60 versus 0.68; incremental *R*^2^ = 0.02 versus 0.05), demonstrating that our feature selection strategy substantially reduced model complexity while retaining comparable predictive performance.

#### Benchmarking results

We benchmarked our PRS*_G_* against PRS derived using alternative published GWAS strategies, including a scaleable mixed model, SAIGE, (PRS_SAIGE_; *p <* 10*^−^*^8^)[31] and two PRS constructed from external European GWAS summary statistics (PRS_LALL_and PRS_SCOTT_)[32–34] (Figure 2) not included in UK Biobank. Thus, despite using only 809 variants, PRS*_G_* achieved comparable risk stratification for T2D (ORs: 1.79 and 1.85 for the top 10% and 20%, respectively) to PRS_SCOTT_ (1.83, 1.95; *∼*360-fold more variants), while outperforming both PRS_LALL_ (1.75, 1.78; 972 variants) and PRS_SAIGE_ (1.64, 1.67; 153 variants). These results establish PRS*_G_* as a robust baseline for evaluating the incremental value of multi-locus encoding.

**Figure 2:**
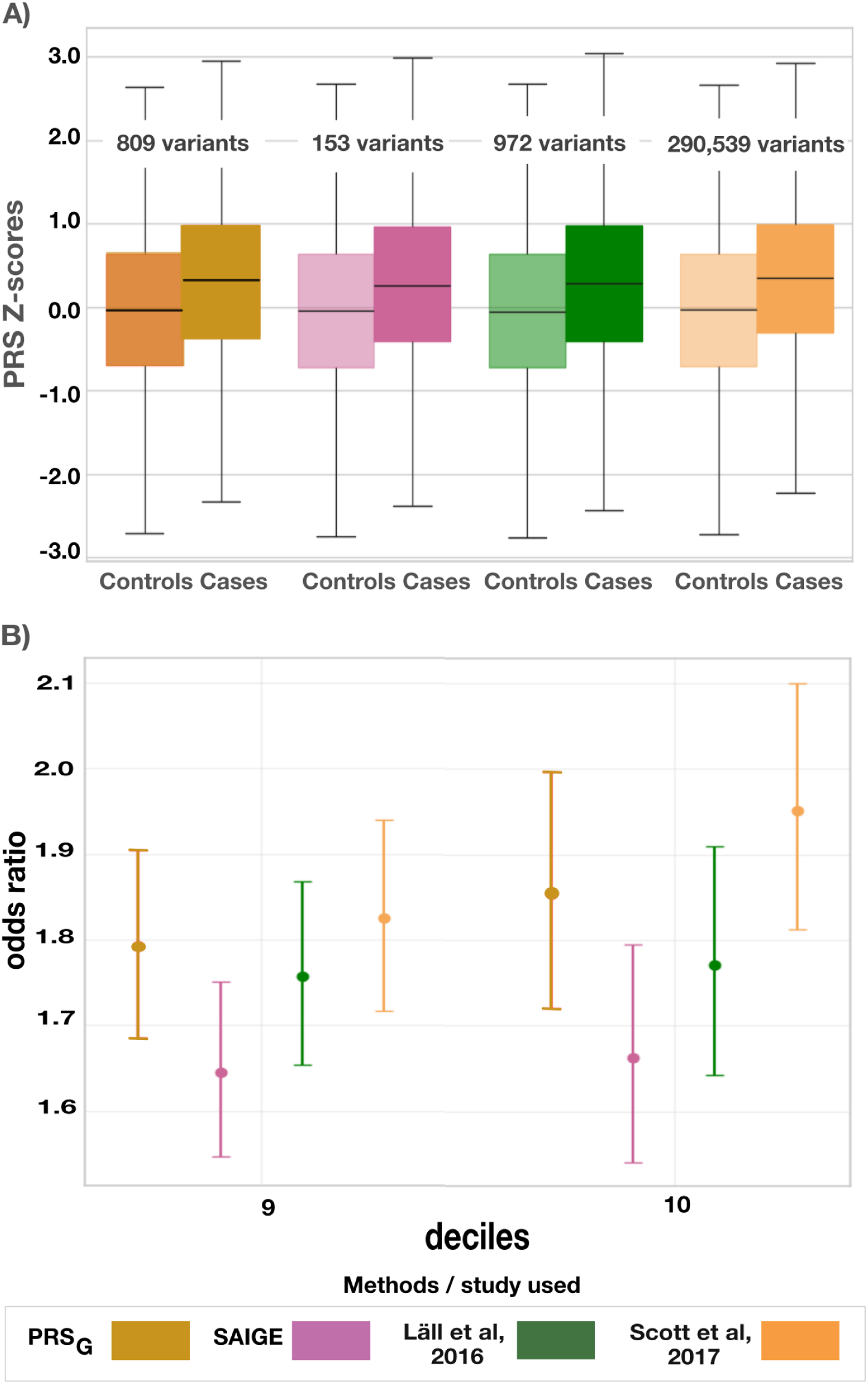
T2D PRS*_G_* benchmarked against published PRS methods and summary statistics. (A) PRS*_G_* performance in discriminating T2D cases and controls was compared to PRS built using alternative GWAS methods (SAIGE) and summary statistics from recent European-ancestry studies (Lall, Scott). The solid lines represent the median and the quantiles are shown as box plots. (B) Odds ratio (OR) and 95% CI are shown for individuals in the top 10% and 20% PRS percentiles across methods. “PRS*_G_*” refers to our current study (mustard hues).

### 2.2 G+G and G*×*G encoding strategies yield complementary, statistically distinct high-risk cohorts

To determine whether multi-locus models captured information distinct from PRS*_G_* and from each other, we assessed pairwise model redundancy using three complementary criteria: (1) Cohen’s *κ <* 0.40, (2) Jaccard index *<* 0.50, and (3) DeLong’s test *p <* 0.05 with |ΔAUC*| >* 0.05. Model pairs failing at least two criteria were considered redundant (Methods 4.7).

Considering the combined models together, no additional redundancy was detected in T2D, and five models were retained. In CD, PRS*_G_*_+*G*_ was redundant with PRS*_G_*, whereas PRS*_GxGxE_* was redundant with PRS*_G_*_+*GxE*_; the latter only was retained because it achieved the larger AUC. Consequently, five models were retained for T2D (PRS*_G_*, PRS*_GxG_*, PRS*_G_*_+*G*_, PRS*_GxGxE_*, PRS*_G_*_+*GxE*_) and three for CD (PRS*_G_*, PRS*_GxG_*, PRS*_G_*_+*GxE*_). Overall, the retained models showed greater distinctness in T2D than CD (Table 1), reflecting differences in genetic architecture. In T2D, G+G and G*×*G remained non-redundant despite being derived from the same SNP pairs, whereas in CD, G+G was redundant with PRS*_G_* while G*×*G and G+G*×*E remained distinct.

### 2.3 Multi-locus encoding strategies identify complementary high-risk cases beyond traditional PRS

Multi-locus ePRS encoding strategies consistently identified high-risk cases not captured by traditional additive PRS in both diseases. Across retained models, 78% (4,187/5,392) of T2D cases and 77% (575/748) of CD cases in the validation set were identified, with 53% and 44% of identified cases exclusive to one model (i.e. missed with every other model), respectively (Table 2). These findings show that aggregate discrimination metrics such as AUC, which evaluate models separately across the entire cohort, can underestimate the value of complementary feature encodings for identifying distinct high-risk individuals (Figure 3).

**Figure 3:**
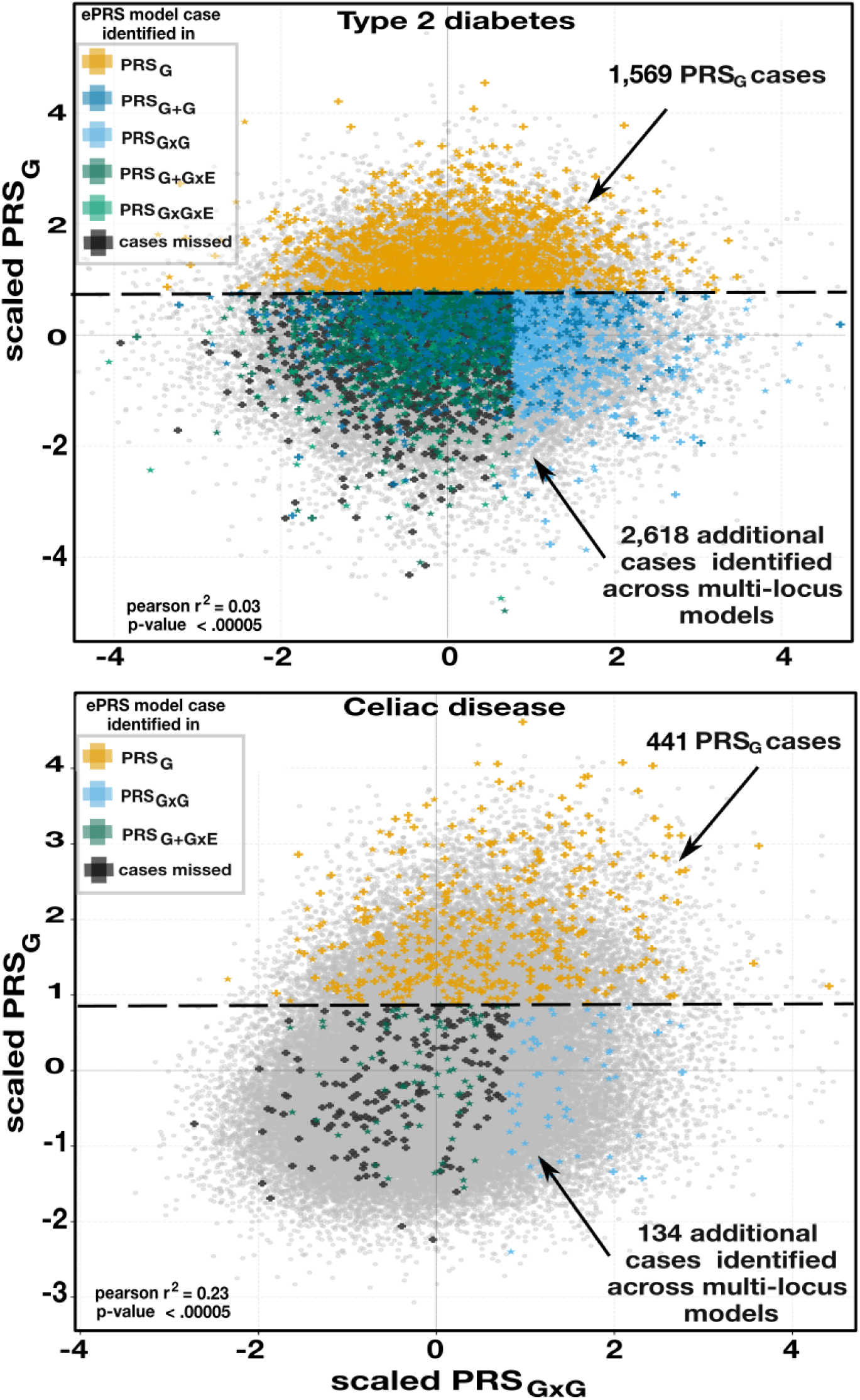
Statistically distinct ePRS models distinguish non-redundant high-risk cases in T2D and CD across feature encoding strategies. Each panel shows scaled individual risk scores for PRS*_G_* (*y*-axis) versus PRS*_GxG_* (*x*-axis) in the validation set (*n* = 67,402). Controls are grey circles; cases are plus symbols, the top 20% at-risk cases are coloured by identifying model in hierarchical order to avoid overlap. The dashed line shows the PRS*_G_* high-risk threshold used. **T2D (top):** Pearson *r*^2^ = 0.03; 4,187/5,392 cases identified across models, including 2,618 missed by PRS*_G_* alone; 2,200 (53%) unique to a single model. **CD (bottom):** Pearson *r*^2^ = 0.23; 575/748 cases identified, including 134 missed by PRS*_G_* alone; 252 (44%) unique to a single model. Total and model-specific counts are reported in Table 2.

#### Type 2 Diabetes

All four multi-locus models captured cases not identified by PRS*_G_*, averaging 61% of its high-risk case count. PRS*_G_*_+*G*_ captured 765 such cases (49%; *κ* = 0.30 vs. PRS*_G_*), including 301 exclusive cases. Despite lower aggregate performance (AUC 0.52 vs. 0.59; incremental *R*^2^ = 0.001 vs. 0.017) and a smaller high-risk case count (1,166 vs. 1,569), PRS*_GxG_* captured 818 such cases, 341 of which were exclusive, and showed near-complete independence from PRS*_G_* (*κ* = 0.01), indicating that retained G*×*G features capture information beyond additive genetic effects rather than predicting the same cases more accurately. Environmental interaction models contributed the largest complementary signal despite using only 17-27 interaction features. PRS*_G_*_+*GxE*_ captured 1,076 such cases (516 exclusive), while PRS*_GxGxE_* captured 1,125 (745 exclusive), demonstrating that features encoding environmental interactions capture a distinct component of disease risk rather than simply amplifying non-linear genetic effects (Tables 1-2).

#### Coeliac Disease

Although three ePRS models (PRS*_G_*, PRS*_GxG_*, PRS*_G_*_+*GxE*_) identified 77% of CD cases, their greater redundancy reflected the disease’s HLA-dominated genetic architecture. Because HLA variants were excluded from the PLINK epistasis screen, these results specifically reflect non-HLA epistasis-informed interactions, supporting an interaction component of CD risk beyond its dominant additive HLA signal. Across the three retained models (PRS*_G_*, PRS*_GxG_*, and PRS*_G_*_+*GxE*_), PRS*_G_* identified the largest number of cases (441; recall=59%), while PRS*_GxG_* and PRS*_G_*_+*GxE*_ identified 60 and 88 additional cases missed with PRS*_G_* respectively. Overall, 134 cases (23%) were missed by PRS*_G_* alone, and 44% of all identified cases were exclusive to just one of the three retained model (Tables 1-2).

#### Cross-phenotype pattern: disease architecture influences, but does not eliminate, complementary risk capture

The extent of complementary risk capture differed between diseases but was evident in both. In T2D, all multi-locus encodings contributed additional high-risk individuals despite similar aggregate performance metrics, with PRS*_GxGxE_*identifying more than twice as many exclusive cases as PRS*_GxG_* (745 vs. 341). In CD, strong additive HLA effects limited the contribution of multi-locus models to identify distinct high-risk subgroups, yet a substantial numbers of unique cases were still identified. Together, these findings show that multi-locus encoding captures distinct components of genetic risk that are not fully represented by traditional additive PRS across diseases with markedly different genetic architectures.

### 2.4 Gene-environment interaction models identify distinct clinical and environmental risk profiles

To evaluate how gene-environment effects contribute in the various risk models, a distinction that aggregate discrimination doesn’t capture, clinical measures were examined both marginally and as the environmental partners of the features retained in gene-environment interaction models (14 such measures in T2D and 37 in CD), across validation and holdout cohorts (Extended Table S7). For T2D, clinical performance was represented by the measures in routine diagnostic use: glucose, HbA1c and BMI (Extended Tables S4, S12). CD has no equivalent quantitative panel, as diagnosis relies on serological detection of anti-transglutaminase and related autoantibodies [38]. We therefore selected clinical measures for CD empirically, retaining those reaching a Shapley *z >* 2 on their own, which identified urea, basal metabolic rate and haemoglobin concentration (Extended Table S12). Baseline environmental measurements collected at the initial assessment preceded diagnosis for most T2D cases (69%), whereas most CD cases had prevalent disease at baseline, indicating that CD environmental profiles should be interpreted in the context of established disease (Table 3).

**Table 3:** Demographic and clinical characteristics of ePRS model high-risk case groups for Type 2 Diabetes (T2D) and Coeliac Disease (CD) in the validation set.

| | Controls | Cases(all) | $G$ | $G \times G$ | $G+G$ | $G \times G \times E$ | $G+G \times E$ |
| --- | --- | --- | --- | --- | --- | --- | --- |
| **Type 2 Diabetes (T2D)** |  |  |  |  |  |  |  |
| $n =$ | 62,010 | 5,392 | 1,569 | 1,166 | 1,555 | 1,574 | 1,539 |
| Male sex (%) | 44.8 | 60.1 | 59.3 | 61.7 | 60.1 | 60.5 | 57.1 |
| Age at assessment | 56.5 | 60.0 | 59.8 | 59.9 | 59.9 | 61.3 | 60.4 |
| T2D diagnosis age | — | 63.4 | 62.7 | 63.4 | 62.9 | 64.6 | 63.4 |
| Diagnosed post-assessment(%) | — | 69.2 | 66.4 | 71.1 | 66.9 | 70.5 | 66.1 |
| **Coeliac Disease (CD)** |  |  |  |  |  |  |  |
| $n =$ | 66,656 | 748 | 441 | 219 | — | — | 325 |
| Male sex (%) | 46.2 | 33.2 | 32.0 | 34.7 | — | — | 36.6 |
| Age at assessment | 56.9 | 57.1 | 57.6 | 57.4 | — | — | 57.1 |
| CD diagnosis age | — | 52.6 | 52.2 | 52.7 | — | — | 52.5 |
| Diagnosed post-assessment(%) | — | 43.3 | 40.4 | 43.8 | — | — | 44.6 |

Unlike the G*×*G and G+G features, which exclude HLA because the epistasis screen omitted the MHC region, environmental interaction features were built from HLA allelotypes and reduced G, G+G, and G*×*G feature sets (Methods 4.2). In CD, HLA allelotypes entered 38 of the 102 retained G+G*×*E encodings (37%), and although no HLA allele was specified as a candidate in advance, those recovered were the alleles of the established coeliac risk haplotype: DQ2.5 (DQA1*05:01, DQB1*02:01) and DQ8 (DQB1*03:02). T2D showed no comparable involvement, with HLA allelotypes entering only 4 of its 44 environmental interaction encodings (9%), as expected for a disease that is not HLA-mediated.

Notably, the environmental partners for CD were anthropometric (quantitative physical measurements of the human body) and haematological measures reflecting malabsorption, and the case-control difference in these measures was larger in risk-allele carriers than non-carriers (hip circumference with DQB1*02:01: *−*0.32 SD against *−*0.06 SD). Because these measurements followed diagnosis in most CD cases, we interpret this as disease consequence rather than causal interaction. It nonetheless suggests a clinical application: as HLA typing is already routine in coeliac diagnosis, genotype may help identify patients at risk of more severe malabsorption who would benefit from closer nutritional monitoring and earlier diagnosis, while non-carrier cases appear to follow a milder course. The corresponding T2D features are less effected by this concern, as exposure measurement preceded diagnosis in most cases.

**Type 2 Diabetes.**

#### 2.4.1 Case-control differences

Relative to controls, T2D cases exhibited higher basal metabolic rate (BMR; +13%), C-reactive protein (CRP; +59%), systolic and diastolic blood pressure, and lymphocyte counts, while LDL cholesterol and the Polyunsaturated Fatty Acids to Monounsaturated Fatty Acids (PUFA:MUFA) ratio were reduced, consistent with obesity, systemic inflammation, and metabolic dysregulation (Table 4).

**Table 4:**
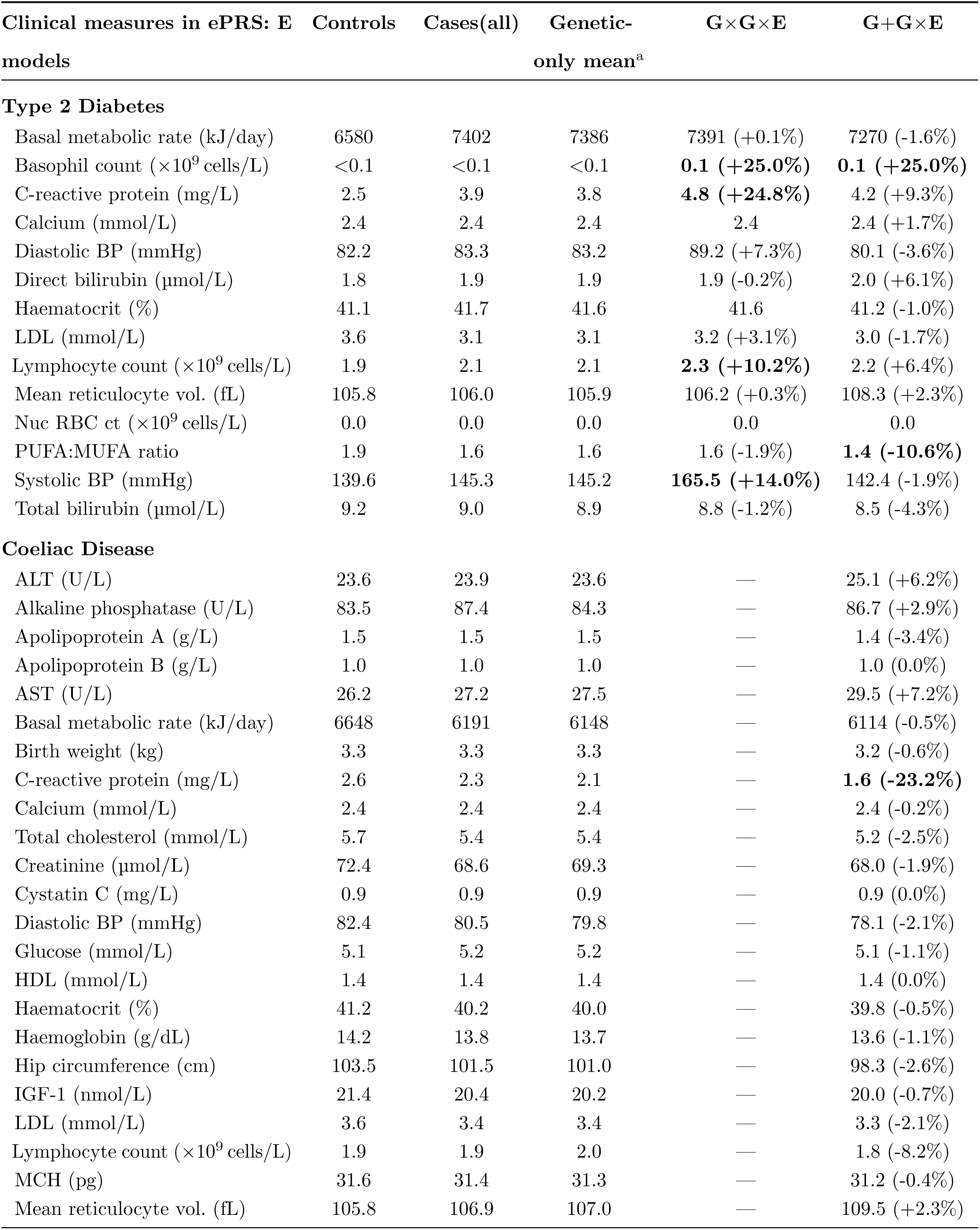
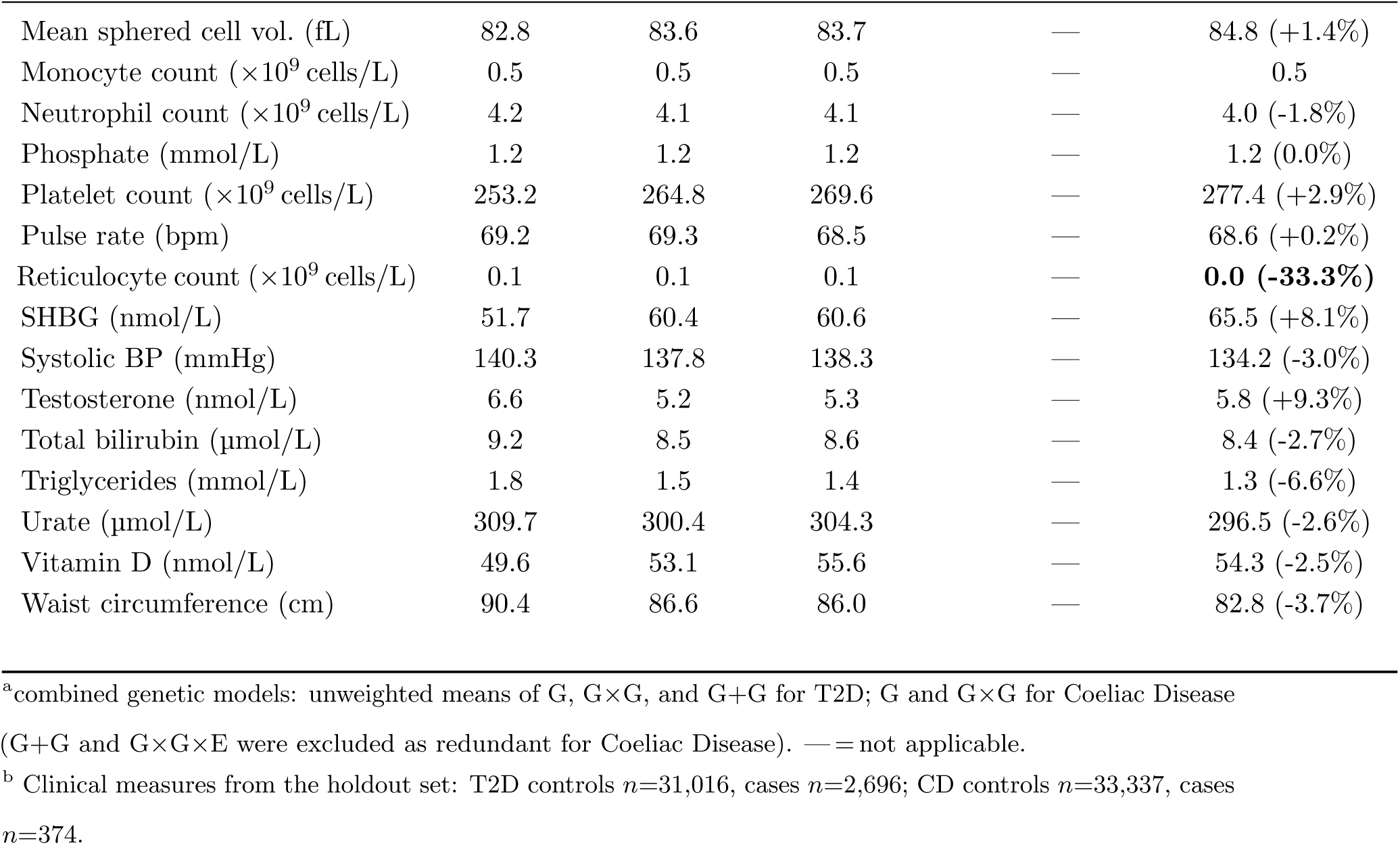
Mean values of clinical and biomarker features in ePRS high-risk groups in the holdout set, with percentage difference from the genetic-only mean shown in parentheses for gene-environment models. Values in bold differ by *≥*10% from the genetic-only mean.

#### 2.4.2 Distinct environmental subtypes identified by interaction models

The two gene-environment interaction models identified markedly different high-risk clinical profiles that replicated in the holdout cohort. The G*×*G*×*E model was characterised by elevated systolic and diastolic blood pressure, increased CRP, and higher lymphocyte counts, consistent with an inflammatory, hypertension-associated subtype. In contrast, the G+G*×*E model showed comparatively normal blood pressure but lower BMR and PUFA:MUFA ratio together with elevated basophil count and mean reticulocyte volume, indicating a distinct metabolic and haematological subtype. The environmental profiles for the G, G*×*G, and G+G models, however were similar to the overall T2D case population (Table 4).

#### 2.4.3 Clinical measures

HbA1c, BMI, and glucose distributions were highly consistent between validation and holdout cohorts. Cases showed the expected elevations relative to controls, while high-risk individuals identified by different ePRS models displayed broadly similar HbA1c, BMI, and glucose distributions, indicating that model-specific differences primarily reflected environmental interaction profiles rather than conventional diagnostic measures (Extended Table S11).

##### Coeliac Disease

Haemoglobin, BMR, and urea distributions were highly reproducible between validation and holdout cohorts (Extended Table S15) with a modest reduction in cases for all three measures (Table 4). The largest departures from the genetic-only profile were in C-reactive protein and reticulocyte count, the only CD measures differing by more than 10% from the genetic-only mean, both lower in the G+G*×*E high-risk group (*−*23.2% and *−*33.3% respectively). Reduced reticulocyte production alongside lower haemoglobin is consistent with a deficiency-limited rather than a compensated anaemia. Alongside a lower BMI, the lower C-reactive protein indicates that this subgroup is distinguished by malabsorption rather than by systemic inflammation. Overall, environmental and clinical differences were modest but consistently replicated across independent cohorts (Table 4).

### 2.5 ePRS feature sets correspond differentially to functional clusters associated with established T2D subtypes

Model-specific ePRS features (Methods 4.8 4.9 4.10) mapped preferentially to distinct T2D functional clusters [39–43], and through them to the disease subtypes [44–47] those clusters have been associated with, consistent with alternative multi-locus encodings capturing complementary disease mechanisms rather than redundant polygenic signals. The correspondence is indirect: features are assigned to published functional clusters, and those clusters carry previously reported subtype annotations. Using SHAP-selected genomic and clinical features from high-risk holdout cohorts, we identified 271 model-specific features, comprising 204 genomic features (340 unique SNPs mapping to 212 loci) and 67 clinical measures, of which approximately 25% mapped to established T2D functional clusters (Extended Tables S8-9; Figure 4). Features spanned three major biological categories: autoimmune, *β*-cell/glycaemic, and metabolic pathways. PRS*_G_*_+*G*_ showed the broadest biological representation and was enriched above chance across both the *β*-cell/glycaemic and metabolic clusters, while PRS*_G_* was most strongly enriched in the obesity clusters. PRS*_GxG_* was concentrated in metabolic clusters, particularly liver/lipid metabolism and obesity, and was the only model contributing features to the HLA-autoimmune cluster. PRS*_GxGxE_* was confined largely to metabolic clusters, whereas PRS*_G_*_+*GxE*_ was distributed across metabolic and *β*-cell dysfunction clusters. These patterns corresponded to established T2D subtypes: PRS*_GxG_* was depleted across subtypes except SAID, where its HLA-associated features reached chance levels, while PRS*_GxGxE_*contributed features to the metabolically driven mild obesity-related diabetes (MOD), mild age-related diabetes (MARD), and severe insulin-resistant diabetes (SIRD) subtypes but none annotated to severe autoimmune diabetes (SAID) or severe insulin-deficient diabetes (SIDD) [44–47] (Extended Tables S8-10).

**Figure 4:**
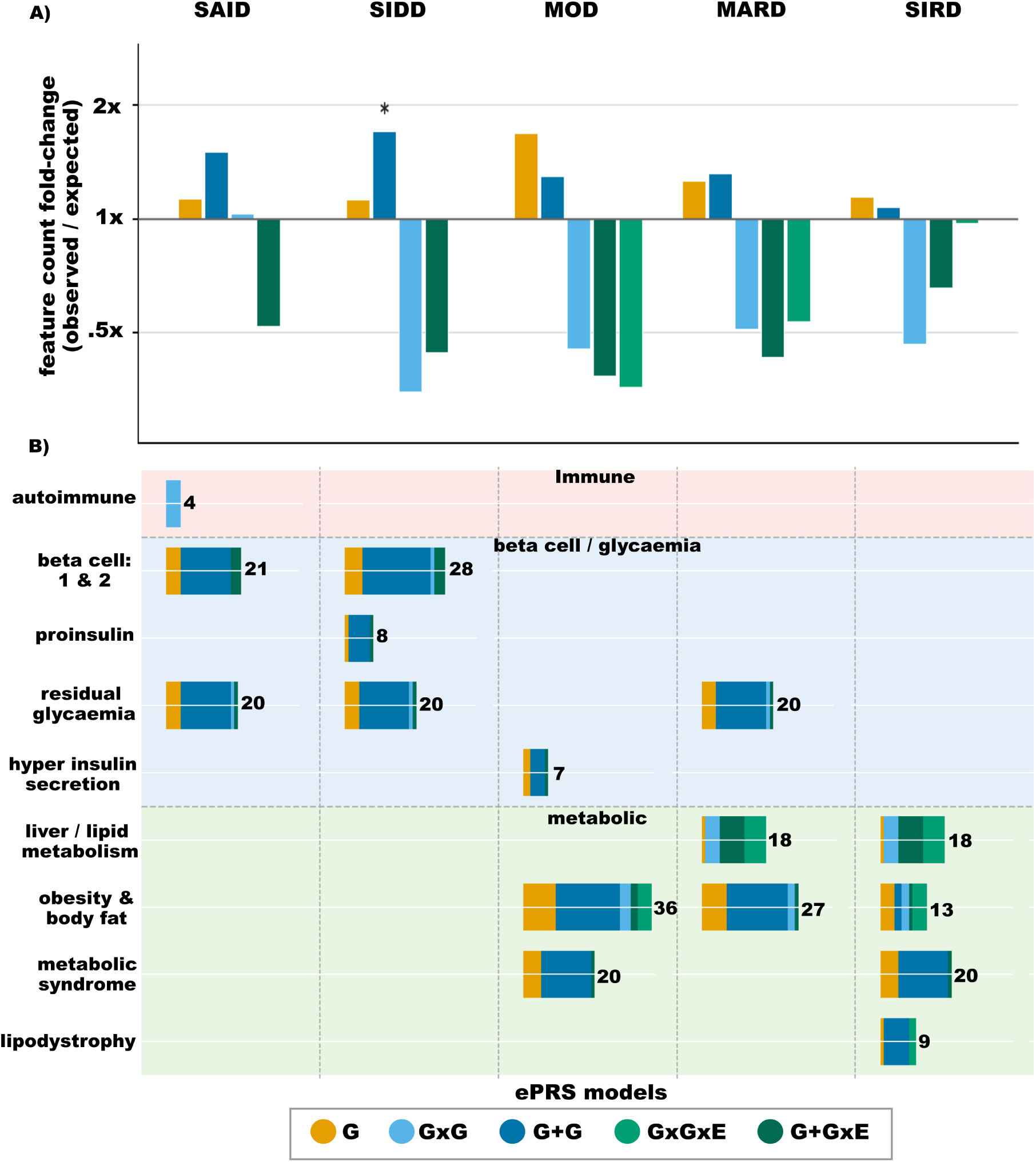
ePRS feature sets correspond differentially to functional clusters associated with T2D subtypes. **A)** Enrichment of PRS models within each T2D subtype, pooled across the clusters associated with that subtype in prior research. Fold enrichment (observed/expected under 10,000 label permutations; is plotted on a log2 scale, equidistant for doubling and halving around the 1*×* baseline; axis labels show the corresponding fold value. Asterisks indicate *q <* 0.05 (Benjamini-Hochberg corrected). **B)** Feature counts by published functional cluster (rows) [39–45, 47] and T2D subtype (columns), on a single linear scale shared across the grid; totals printed to the right of each bar. Dashed lines separate pathway groups (Immune, *β*-cell/glycaemia, Metabolic). Colours denote PRS model throughout. SAID=severe autoimmune diabetes, SIDD=severe insulin deficient diabetes, SIRD=severe insulin resistant diabetes, MOD=mild obesity related diabetes, MARD=mild age-related diabetes. Panels need not agree: **A** pools across clusters per subtype and corrects for model size; **B** keeps clusters separate and shows uncorrected counts.

Permutation testing provided formal support for two model-specific associations. Among 75 annotated features, models contributed 11-27 each, with one of 43 model-by-cluster cells and one of 23 model-by-subtype cells reaching *q <* 0.05. PRS*_GxG_* was enriched in the HLA-autoimmune cluster (4.0 observed vs. 0.6 expected; *q* = 0.034), while PRS*_G_*_+*G*_ was enriched in SIDD (24.0 vs. 14.1; *q* = 0.005). All directionally annotated features agreed with their cluster’s expected direction, with no net discordance across models. Given the small number of annotated features per model, these tests were conservative; for example, PRS*_G_*_+*G*_ contributed 18 features to the *β*-cell cluster versus 10.2 expected, but did not reach significance (*q* = 0.073).

Together, these findings show that the feature sets selected by our alternative ePRS encodings differ in subtype annotations: PRS*_GxGxE_* features fall only in the metabolically driven MOD, MARD and SIRD subtypes and none in clusters annotated to SAID or SIDD, while the autoimmune cluster is annotated exclusively by PRS*_GxG_* features. Enrichment above chance was nonetheless confined to PRS*_G_* and PRS*_G_*_+*G*_, and only PRS*_G_*_+*G*_ in SIDD survived correction for multiple testing. This is an association between feature sets and previously published cluster annotations rather than a direct partitioning of patients into molecular subtypes, and because power is bounded by the number of annotated features per model, the absence of enrichment among the interaction encodings is not evidence that they lack subtype correspondence. The differing composition is suggestive of complementary disease mechanisms beyond traditional additive polygenic risk scores, but this analysis does not establish it.

### 2.6 Composite ePRS enhances risk prediction beyond PRS*_G_*

If individual ePRS models capture complementary risk, combining them should improve prediction. We therefore constructed ePRS_Comp_ by fitting a single logistic regression over the retained ePRS models in the validation set and applying it once to the untouched holdout set (Methods 4.11). To test whether weighting the models matters, we compared it with a rule-based union, ePRS_Rule_, which flags anyone placed in the top 20% by any retained model (Methods 4.11).

In the holdout cohort, ePRS_Comp_ improved T2D discrimination over PRS*_G_* (AUC 0.65 vs. 0.58) and more than doubled risk stratification in the top decile (OR 6.5 [95% CI: 5.2–8.2] vs. 2.5 [2.1–3.0]; Figure 5). All five encodings carried an independent weight, and the two G*×*E encodings carried the largest (PRS*_GxGxE_ β* = 0.30, PRS*_G_*_+*GxE*_ *β* = 0.28, against PRS*_G_ β* = 0.21; all *p <* 0.01; Extended Table S3), direct evidence that the encodings capture partially independent risk. At the 64.4% coverage of ePRS_Rule_, ePRS_Comp_ identified 2,179 of 2,696 cases (80.8%), against 2,077 (77.0%) for ePRS_Rule_ and 1,990 (73.8%) for PRS*_G_*; at a 20% budget it identified 965 cases against 763 for PRS*_G_*. ePRS_Rule_ also reached a lower top-decile OR (4.1 [3.3–5.1]) and AUC (0.61).

**Figure 5:**
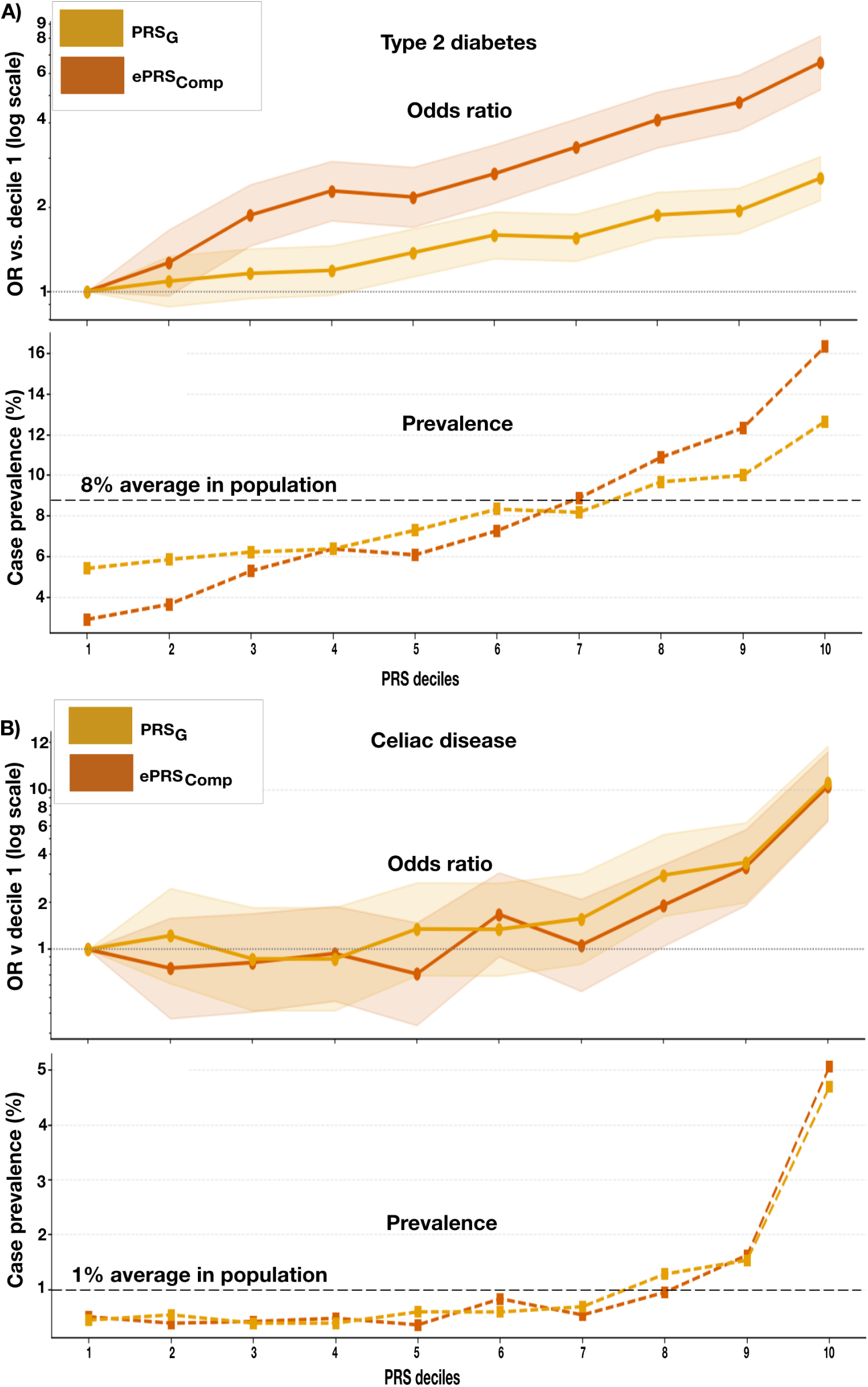
ePRS composite score improves risk stratification over PRS*_G_* in T2D, with limited gain in CD. Each panel shows risk stratification for ePRS_Comp_ (dark mustard) and PRS*_G_* alone (light mustard) across deciles 1-10 in the holdout set. Top row in each panel shows odds ratio relative to decile 1 (OR = 1) (*y*-axis, log scale) with 95 % CI as shaded ribbons; the bottom row of each panel shows case prevalence (*y*-axis, %) relative to population average (dashed black line). **(A) T2D:** ePRS_Comp_ achieves OR 6.5 (95 % CI: 5.2–8.2) in the top decile (prevalence 16%) versus 2.5 (95 % CI: 2.1–3.0) and 13% for PRS*_G_* alone. **(B) CD:** ePRS_Comp_ and PRS*_G_* perform similarly in the top decile: OR 10.5 (95 % CI: 6.4–17.4) versus 11.1 (95 % CI: 6.5–18.9), prevalences 4% and 5%. Wider CIs relative to T2D reflect the *∼*5-fold lower disease prevalence and sparse case counts per decile.

In CD, combining models gave essentially no gain. ePRS_Comp_ reached an AUC of 0.75 against 0.74 for PRS*_G_*, with no improvement in top-decile stratification (OR 10.5 [6.4–17.4] vs. 11.1 [6.5–18.9]), and at the 45% coverage of ePRS_Rule_ it identified 293 of 374 cases against 283 for both PRS*_G_*and ePRS_Rule_. The fitted weights show why: PRS*_G_* carried almost all of the signal (*β* = 0.84), PRS*_GxG_* contributed nothing detectable (*β* = 0.02, *p* = 0.58), and only PRS*_G_*_+*GxE*_ added modestly (*β* = 0.27), consistent with CD’s HLA-dominated architecture[48, 49]. ePRS_Rule_ reached an AUC of 0.72, no better than PRS*_G_* (*p* = 0.11), because a union of scores dilutes the score when only one encoding is informative.

In both diseases the fitted ensemble outperformed the rule-based union (DeLong *p* = 3.7 *×* 10*^−^*^28^ in T2D; *p* = 2.7 *×* 10*^−^*^3^ in CD), and the contrast between architectures is visible in the weights themselves: every encoding contributes in T2D, whereas in CD the interactive encoding is indistinguishable from zero. Together, these findings support the hypothesis that complementary multi-locus encodings capture partially independent components of genetic risk, and that weighting them, rather than taking their union, is what converts that complementarity into improved prediction.

#### 2.6.1 Composite ePRS provides complementary information to clinical measures

We next asked whether ePRS_Comp_ retains risk discrimination among individuals that conventional clinical measures do not flag (Figure 6; Extended Tables S5-6). Because HbA1c, BMI, and glucose are established diagnostic measures for T2D, we defined subgroups by individuals not exceeding the clinical threshold for each measure individually, and for all three combined. Within each subgroup, precision, the fraction of correctly identified cases in a given score decile, is plotted across deciles 10 (highest score) to 1 for both ePRS_Comp_ and PRS*_G_*, with decile boundaries computed separately for each score within each subgroup (Methods 4.11).

**Figure 6:**
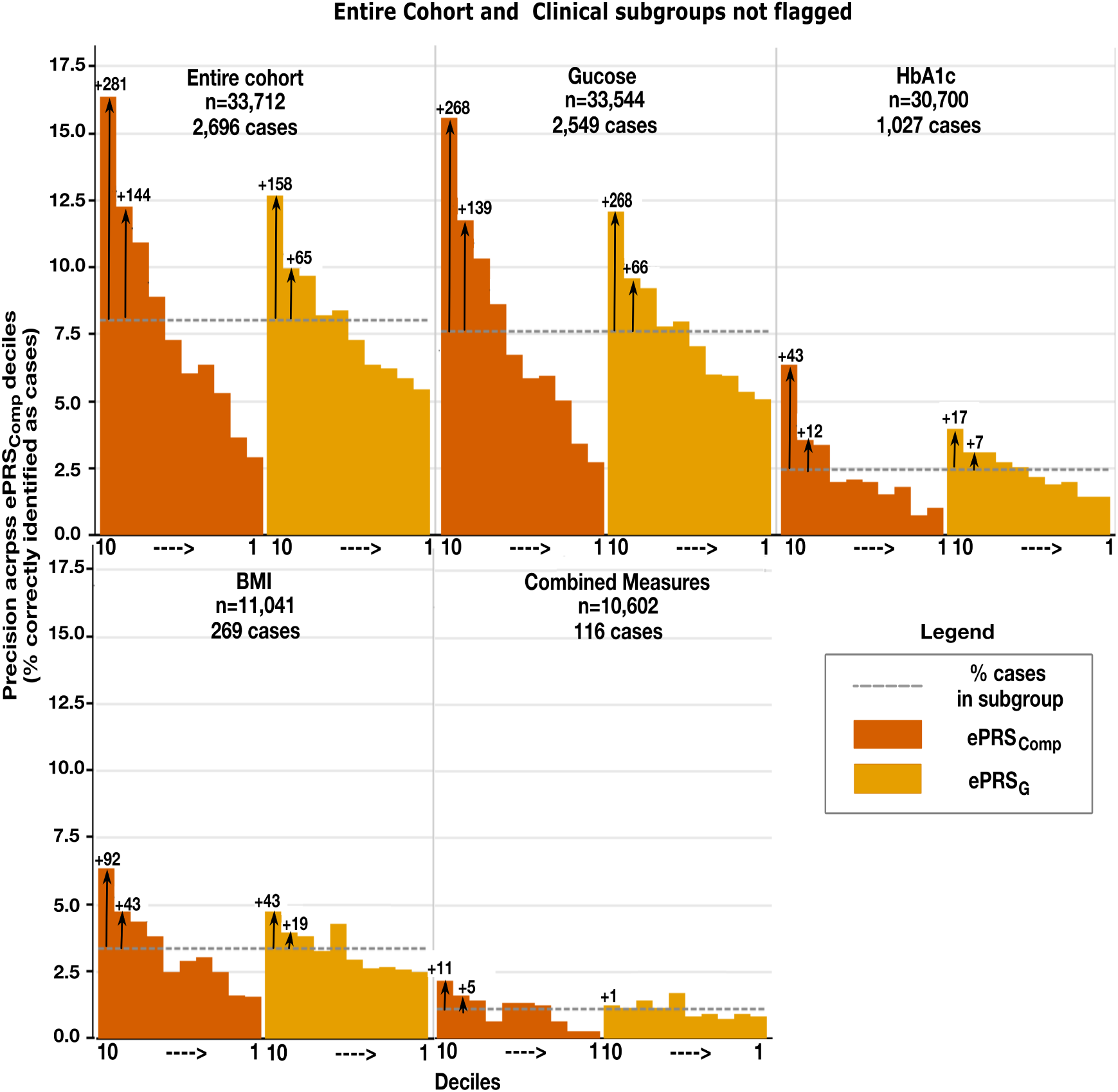
ePRS_Comp_ concentrates T2D cases among individuals not flagged as high-risk with clinical measures. Subgroups comprise individuals not identified as high risk by the clinical measure named (subgroups), the full cohort (ePRS_Comp_ only; top left) is shown for reference. Subgroups are ordered by case rate, with size and case count annotated above each group. Bars show precision across descending ePRS deciles (x-axis); dashed lines indicate subgroup case rates, with the difference representing enrichment over random identification. Arrows and counts indicate additional cases identified by scores in deciles 9–10 above prevalence. Not flagged” indicates values below the threshold for HbA1c (41 mmol/mol), BMI (25 kg/m^2^) or glucose (12 mmol/L); ”Combined Measures” requires all three below threshold. Coeliac disease (CD) is omitted because ePRS_Comp_ offers no advantage over PRS*_G_*.

ePRS_Comp_ retained discrimination in clinically unflagged subgroups across all three measures (Figure 6). In the full holdout cohort, the top 20% of ePRS_Comp_ (deciles 9–10) captured 965 cases (14.3% precision) versus 763 (11.3%) for PRS*_G_* (AUC 0.65 vs. 0.58). Precision remained above the subgroup case rate for both scores in every subgroup examined. Enrichment was strongest among individuals with BMI below 25 kg/m^2^, where ePRS_Comp_ captured 109 cases (4.9%) versus 78 (3.5%) for PRS*_G_* (AUC 0.67 vs. 0.59, *p* = 3.7 *×* 10*^−^*^6^), and remained significant among those not flagged by HbA1c (341 vs. 268 cases; 5.6% vs. 4.4%; AUC 0.62 vs. 0.56, *p* = 1.5 *×* 10*^−^*^12^). Enrichment was weakest among individuals not flagged by any of the three measures (10,602 individuals; 116 cases; 1.09% prevalence), where the top 20% of ePRS_Comp_ captured 40 cases (1.9% precision; 1.7-fold enrichment) versus 25 cases (1.2%) for PRS*_G_* (AUC 0.63 vs. 0.55, *p* = 8.7 *×* 10*^−^*^4^). The modest AUC decline from the full cohort (0.65) indicates that ePRS_Comp_ captures genetic risk beyond clinically informative measures, though its incremental contribution is limited by the small size and low prevalence of the most restrictively defined subgroups.

Importantly, the gene-environment model comparison is independent of clinical measures presented here: none of HbA1c, BMI or glucose is an input to the 14 clinical measures entering gene-environment features retained in T2D, therefore ePRS_Comp_ does not encode the clinical information it is compared against. These findings indicate that ePRS_Comp_ retains discrimination where conventional clinical measurements fail. However, they do not establish clinical utility. Combined clinical measure thresholds flag 69% of the cohort and capture 96% of all cases (2,580 of 2,696). Among the 10,602 individuals not flagged by any measure, the top 20% of ePRS_Comp_ captures 40 of the 116 cases otherwise missed, with 16 cases above the subgroup prevalence; PRS*_G_* recovers 25 cases with only 1 above the subgroup prevalence, essentially at subgroup baseline. The yield is real but small. Whether a composite score can be calibrated to a classification budget at which this becomes worthwhile remains to be determined.

In CD, where no diagnostic measure exists, the corresponding comparison used the three clinical measures selected empirically (basal metabolic rate, urea and haemoglobin concentration). Together these reached an AUC of 0.62, rising to 0.72 when ePRS_Comp_ was added, and each measure gained comparably on its own (0.59-0.60 alone, 0.72 with ePRS_Comp_; Extended Table S5). The increment is far larger than in T2D (ΔAUC *∼* 0), where the clinical measures are themselves diagnostic criteria.

## 3 Discussion

### Epistasis-informed and multi-locus feature encoding identifies non-overlapping high-risk individuals beyond additive PRS, offering a window into missing heritability

Traditional PRS aggregates marginal SNP effects under an additive assumption, yet the gap between estimated SNP heritability and variance explained by current PRS (i.e. missing heritability [50]) has multiple sources [5, 8, 51], including non-additive genetic effects that standard GWAS systematically excludes. Our results are consistent with the hypothesis that this exclusion is consequential and under estimates true PRS. In T2D, five statistically distinct ePRS models identified 4,187 cases in our present study (78%) in the top-20% risk group, with 2,200 cases (53%) uniquely captured by a single model missed by all others, including PRS*_G_*. Each feature encoding strategy identified hundreds of exclusive high-risk cases that were missed by all other ePRS models (Table 2), with almost no overlap between some models (Table 1), providing further support that distinct biological components discoverable with distinct non-linear feature coding strategies improve PRS calculations. Notably, PRS*_GxGxE_* captured more than twice the exclusive cases of PRS*_GxG_*, consistent with the hypothesis that gene-environment interactions play a significant role in metabolic disease [10, 11]. Notably, standard performance metrics such as AUC, precision, recall, and OR incompletely characterise multi-locus PRS when complementary models identify distinct high-risk individuals despite similar aggregate performance. The same limitation applies when comparing genetic with demographic predictors. In T2D, a covariate-only model of age, sex, and ancestry principal components achieves an AUC of 0.65, exceeding or matching individual ePRS evaluated here confirming that age and sex are strong determinants of T2D. Whereas in CD the same covariates reach an AUC of only 0.58 and are outperformed by PRS*_G_*, with differences reflecting disease architecture rather than model quality. Thus, aggregate discrimination reflects how much population-level risk is explained by readily available predictors, and not if a genetic model identifies individuals missed by those predictors. Here, we distinguish ePRS models based on the individuals they identify and the mechanisms underlying that complementary risk capture.

### The divergence between T2D and CD shows that genetic architecture driving risk determines both optimal feature encoding strategy and the limits of multi-locus interaction models

Applying the ePRS pipeline across diseases with contrasting genetic architectures reveals how risk is partitioned differently across complex diseases. In T2D, these complementary feature encodings yield broadly similar performance, with gene-environment models identifying substantially more unique cases, indicating that environmental modulation exposes additional genetic risk not captured by additive or purely genetic models. In contrast, CD is dominated by large-effect HLA loci, limiting gains from interaction modelling. PRS*_G_* already approaches maximal performance, and PRS*_GxG_* provides modest and largely HLA-independent signal (AUC = 0.58) from non-HLA pairs despite HLA loci being included in ePRS models [9]. The most informative interaction-encoded signal arises from PRS*_G_*_+*GxE*_, where multiple HLA loci enter gene–environment encodings, suggesting that a portion of HLA-mediated risk may be environmentally modulated. Together, these results indicate that the benefit of multi-locus models scales with genetic heterogeneity, with greater measurable gains in more polygenic architectures.

### Integrating complementary feature encodings increases case identification, but the value measured for any score depends on the clinical benchmark it is placed against

Because each of our ePRS models identified partially non-overlapping high-risk individuals, no single feature encoding captured the full spectrum of inherited risk. Applying a modified multiplexed polygenic framework of Patel *et al.* [49], ePRS_Comp_ integrated complementary ePRS models and improved T2D discrimination, capturing genetically high-risk individuals below conventional clinical thresholds. The value of that gain depends on the clinical diagnostics it is measured against, which differed markedly between the two diseases. In T2D, where HbA1c, BMI and glucose are themselves diagnostic criteria, commonly used thresholds for these measures flag 69% of the cohort and capture 96% of all cases; and ePRS_Comp_ does not improve discrimination when added to a fitted clinical model (AUC 0.91 with and without it; Extended Table S5). Its contribution is confined to 10,602 individuals not flagged by any threshold, and is bought at a cost: the top 20% of ePRS_Comp_ among them recovers 40 of the 116 missed cases, requiring 6% of the full cohort to be additionally flagged at roughly 53 individuals per case. Restricting to cases diagnosed at least six years after assessment does not change this, as baseline clinical measures remain prognostic for future diabetes (AUC 0.91 against 0.65 for ePRS_Comp_).

In CD, the genetic score outperformed the available clinical measures, but these were non-specific markers of established malabsorption rather than diagnostic tests; compared with coeliac serology, the incremental value would likely be smaller. Thus, differences between diseases reflect what the available clinical measures capture rather than differences between genetic models, and do not imply that genetic scores should replace established diagnostic testing. More broadly, the value of combining feature encodings depends on the diversity of distinct genetic signals underlying disease, while the apparent benefit of a genetic score depends on the benchmark used. Whether genetic risk adds meaningful information before clinical measurements are available, such as from birth, remains untested here.

### The statistically distinct risk cohorts identified across ePRS models provide a framework for understanding biological heterogeneity in complex disease

These cohorts reflect distinct genetic and genetic–environmental signals captured by the underlying encoding strategies rather than differences in risk score alone. The large number of selected SNP pairs showing departure from additivity provides evidence that these encodings capture relationships beyond simple additive effects, while not establishing functional interaction between the corresponding gene products.

### 3.1 Future Directions

Future work should evaluate these models across independent cohorts and ancestries, improve integration of complementary encodings, and extend T2D subtype frameworks to incorporate interaction-derived features. The interpretable framework also enables SNP pairs, environmental variables, and effect directions to be mapped to pathways and tissues and investigated longitudinally, while the many features showing departure from additivity provide candidates for functional validation.

### 3.2 Limitations

Several limitations should be considered. First, analyses were restricted to European-ancestry participants from UK Biobank, requiring validation in independent cohorts and diverse ancestries. Second, departure from additivity was confirmed for many of the retained G*×*G and G+G pairs, but these tests used the same cohort as feature selection and therefore require replication; effect sizes are less certain, with median *|J |* declining as the number of double carriers increases, suggesting upward bias for less common pairs. Moreover, a third of CD pairs could not be tested under the double-carrier threshold, limiting direct comparison with T2D. Third, none of the T2D and all but 10 of the CD gene–environment features were individually confirmed as statistical interactions and are therefore described as encodings. In CD, these features were dominated by HLA-containing terms whose environmental partners largely reflected post-diagnostic malabsorption, so they are interpreted as disease consequences rather than G*×*E effects. Functional validation is needed to establish biological mechanisms, particularly because HLA variants were excluded from the initial epistasis screen. Finally, biological interpretation was constrained by T2D subtype frameworks based on single-variant associations; incorporating interaction-derived features may better capture the biological heterogeneity identified by ePRS.

## 4 Methods

### 4.1 Overview of this PRS study that includes gene and environmental interactions

We developed novel methods to incorporate additive (G), interaction-screen-informed multilocus additive (G+G), epistasis-screened (G*×*G), and their environmental interaction counterparts (G+G*×*E and G*×*G*×*E) as feature weights into polygenic risk calculations (Figure 1). G*×*G pairwise interactions were identified by exhaustive SNP-SNP interaction analysis on the cleaned genotyped (G) dataset. To reduce noise, an embedded feature selection approach[52, 53] was applied separately to G, G+G, and G*×*G features: a *gradient-boosted classifier* [54] first filtered features by predictive performance in batches of 3K, followed by ranking via Shapley value z-scores (Figure 1B). Significant SNP pairs were then encoded using two strategies: interaction-screen-informed multilocus additive burden (sum of allele counts, G+G) and epistasis-screened features (product of allele counts, G*×*G). Cardiometabolic clinical markers (“E”) with *<* 5% missingness from UK Biobank health records were combined with reduced feature set (G, G*×*G, and G+G*×*G) in G+G*×*E and G*×*G*×*E interaction feature discovery. Seven separate L1-penalized logistic regression models were trained on the reduced feature sets (Figure 1C), and corresponding ePRS scores were computed for the validation set using non-zero *β* coefficients; statistically distinct models were retained for downstream analysis (Figure 1D).

### 4.2 Data sources

We used the UK Biobank, a prospective cohort of 500,000 individuals aged 40-69 at recruitment[28]. Genotyped SNPs, imputed HLA allelotypes, and electronic health record phenotypes were obtained from questionnaires, interviews, and linked clinical records.

#### 4.2.1 Genotyped data

To increase the power of the study to find higher order interactions and consistent with previous benchmarking studies[30] we used a subset of 805,426 genotyped markers in 337,070 individuals of white british ancestry. Inclusion criteria were: (1) self-reported White British ancestry, (2) used in principal component analysis (PCA) performed by UK Biobank, (3) not heterozygosity outliers, (4) no putative sex chromosome aneuploidy, and (5) *≤*10 third-degree relatives, as these can bias GWAS results[55].

#### 4.2.2 Clinical Measures : E

Environmental (E) features for gene-environment interactions were selected from 83 cardiometabolic measures in electronic health records (Extended Table S12). Features with *>* 5% missingness were excluded, resulting in 72 measures included in G+G*×*E and G*×*G*×*E analysis (Extended Tables S12 & S7).

#### 4.2.3 Data cleaning

Genotyped data for phenotypes were independently split into 70% training, 20% validation, and 10% holdout sets each. SNPs were filtered using PLINK v1.9[22] with the following thresholds: missingness *>* 5%, Hardy–Weinberg p-value *<* 5 *×* 10*^−^*^5^, minor allele frequency *<* 10*^−^*^5^, and exclusion of the MHC region (hg19 chr6:25,477,797–36,448,354bp) due to high linkage disequilibrium (LD)[27], and 362 imputed HLA alleloptypes. Post-filtering, 624,035 and 624,034 SNPs remained for T2D and CD, respectively. Additionally, 181 of 362 imputed HLA loci (allele frequency *>* 0.001) were included after converting imputation scores to allele counts (0: *<* 0.7; 1: 0.7–1.7; 2: *≥* 1.7). The final genotyped dataset comprised 337,070 individuals *×* 624,035 SNPs.

#### 4.2.4 Data processing

Cleaned PLINK .bed files were merged and converted to space-delimited .raw format for input into feature selection and modelling pipelines. Covariates included in all models were age, sex, and the first 10 principal components (UK Biobank field 22009, to control for population ancestry).

### 4.3 Phenotype Definitions

We identified individuals with T2D in the DNA Nexus RAP platform with T2D phenotype filters specified in Table 5 and CD phenotype filters specified in Table 6, respectively based on a combination of ICD-10 diagnosis codes, self-reported non-cancer illness codes, confirmed reporting dates for “E11” for T2D, and source of report of K90 for CD, resulting in a dataset with 8% prevalence of T2D and 1% prevalence for CD.

**Table 5:** Phenotype definitions.

| Column | Filter |
| --- | --- |
| Diagnoses - main ICD10 | contains “E11” |
| *Source of report of E11 (non-insulin-dependent diabetes mellitus)* | not blank |
| Non-cancer illness code, self-reported — * (*instances 0-3) | contains “type 2 diabetes” |
| *Date E11 first reported (non-insulin-dependent diabetes mellitus)* | not blank |

**Table 6:** CD phenotype definition.

| Column | Filter |
| --- | --- |
| Diagnoses – main ICD10 | Contains “K90” |
| *Source of report of K90 (intestinal malabsorption)* | not blank |
| Non-cancer illness code, self-reported * (*instances 0-3) | contains “coeliac disease” |

### 4.4 Feature encoding (G, G+G, G*×*G, G*×*G*×*E, G+G*×*E)

#### 4.4.1 Discovery of epistasis-informed interactions (G*×*G pairs)

Pairwise SNP interactions were initially identified using a fully exhaustive search with PLINK v1.9 “fast-epistasis” with the “–boost” modifier[22]. This approach performs case-control association tests for epistatic interactions using log-linear models[12], with the test statistic following a chi-square distribution with 4 degrees of freedom under the null hypothesis of no interaction. A total of 3.9 *×* 10^11^ SNP pairs were analyzed using a default p-value threshold of 5 *×* 10*^−^*^6^ for initial discovery. Analyses were executed on a 40-core node with 30 GB RAM in parallel, requiring *∼*60 hours of wall-clock time. The “–boost” modifier accelerates computation via optimized algorithms while maintaining statistical validity, making it suitable for high-throughput initial discovery prior to feature selection.

#### 4.4.2 Construction of multi-locus features: G+G and G*×*G encoding strategies

Significant SNP pairs from the exhaustive pairwise search were transformed into multi-locus features using two distinct encoding strategies, both applied to the same set of retained pairs following feature selection and ranking.

##### Multilocus additive burden encoding (G+G)

For each significant SNP pair, a burden feature is computed by summing the allele counts across the two loci for each individual:

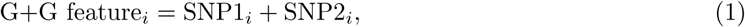

where *i* indexes the epistatic pair and the sum is taken for each individual. This encoding captures the total allelic load across the pair, with values ranging from 0 to 4 (for biallelic loci). Higher values indicate cumulative multi-locus burden regardless of the specific combination of alleles. These features were included in downstream feature selection and ranking for polygenic risk score modelling as PRS*_G_*_+*G*_ and PRS*_G_*_+(*G*+*G*)_.

##### Epistatic product encoding (G*×*G)

For each significant SNP pair, an epistatic interaction feature is computed by taking the product of allele counts across the two loci:

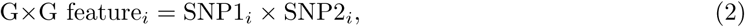

where *i* indexes the epistatic pair and the product is taken for each individual. This encoding captures the statistical interaction between the two loci. The product encoding is non-zero only when both alleles are present simultaneously (values: 0 for single-locus carriers, 1-4 for co-carriers), thereby specifically weighting individuals who co-inherit risk alleles at both loci. This corresponds to the classical definition of statistical epistasis as departure from additive independence[8]. These features were included in downstream modelling as PRS*_GxG_* and PRS*_G_*_+(*GxG*)_.

A direct consequence of these encodings is that while G+G sums over the full range of genotype states with approximately equal weight per allele, G*×*G assigns zero weight to single-locus carriers and non-zero weight only to double carriers.

#### 4.4.3 Construction of environmental interaction features: G+G*×*E and G*×*G*×*E

To facilitate interpretable prediction results, environmental interaction features were created by combining environmental (E) features with the reduced genetic features under each encoding strategy, producing feature sets G+G*×*E and G*×*G*×*E. Centered environmental variables were multiplied in both feature sets with important biological interpretation worth mentioning: G+G*×*E interactions weight the total allelic load at a pair of loci against an environmental variable, whereas G*×*G*×*E interactions weight the specific co-occurrence of alleles at both loci against an environmental variable, a three-way synergistic term. Both feature sets were constructed using identical centering, standardization, and training/validation/holdout split procedures as described below.

E features were first imputed with the training-set mean and mean-centered; the same centering was applied to validation and holdout sets. Interaction terms were then computed by multiplying centered E features with the corresponding encoded genetic features (G+G or G*×*G values). Resulting interaction features were standardized using the training-set mean and standard deviation, with the same parameters applied to validation and holdout sets.

Environmental (E) feature means were calculated from the training set:

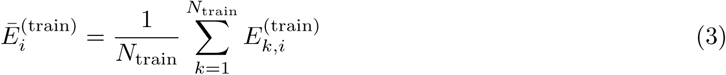

and used to mean-center E features:

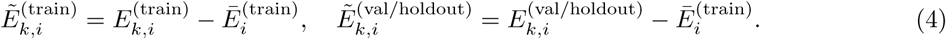

For G+G*×*E features:

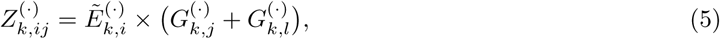

and for G*×*G*×*E features:

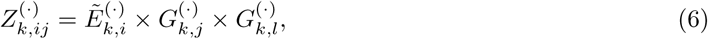

where (*·*) indicates training, validation, or holdout sets, *k* indexes individuals, *i* environmental features, *j* and *l* the two loci in the SNP pair, and *N*_train_ is the number of training samples.

Scaling parameters were computed from the training set:

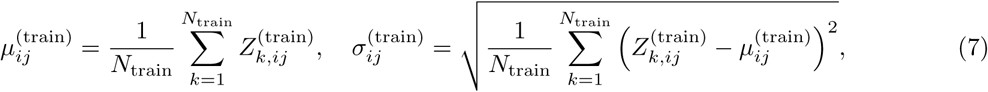

and interaction features (both G+G*×*E and G*×*G*×*E) were standardized:

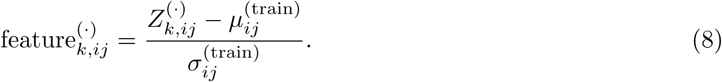

### 4.5 Feature selection and ranking

Dimensionality reduction was performed to reduce computational burden and retain the most informative features for ePRS modelling. Feature selection (FS) included classification, clustering, and regression algorithms, followed by ranking using Shapley feature importance[56, 57].

#### 4.5.1 G, G+G, G*×*G feature selection

A computationally intensive pipeline was implemented to select G, G+G, G*×*G features. Models were trained in batches of 3,000 features to reduce memory requirements. For each batch, gradient boosted histogram (GBH) and complement näıve Bayes classifiers[58] were trained for T2D and CD. Hyperparameters for GBH models were optimized via three-fold cross-validation and grid search across maximum iterations (1000, 1500, 2000), learning rates (0.001, 0.01, 0.1), and L2 regularization penalties (0, 0.1, 0.5) to prevent overfitting[59].

In total, 208 G and 202 G*×*G and G+G models were trained on a high-performance node (50 Intel CPUs, 700 GB RAM, *∼*5 h per model). Models achieving AUC *>* 0.51 were retained; features in lower-performing models were excluded from downstream analysis. Selected models were saved in .pkl format for feature ranking. The retained G, G*×*G. and G+G pairs were subsequently used in G+G*×*E and G*×*G*×*E feature discovery respectively and L1-penalized modelling, as described in the feature encoding section above.

The feature selection output for G, G+G, and G*×*G included: 915 G, 1,497 G+G, and 13,767 G*×*G for T2D, and 191 G, 778 G+G, and 4,747 G*×*G for CD.

#### 4.5.2 Linkage disequilibrium (LD) analysis

The post-discovery LD filter removes any pairs whose constituent SNPs are sufficiently correlated to be considered proxies of the same additive signal; pairs surviving this filter therefore represent loci whose joint signal cannot be attributed to correlated tagging of a single underlying variant.

LD pruning was performed post-feature reduction and gene-environment interaction discovery but prior to regression modelling to enhance PRS robustness[60]. The post-discovery LD filter removes any pairs whose constituent SNPs are sufficiently correlated to be considered proxies of the same additive signal; pairs surviving this filter therefore represent loci whose joint signal cannot be attributed to correlated tagging of a single underlying variant. Pairwise LD was computed using PLINK v1.9 with 100 kb windows and a 1 bp slide, and all SNP pairs with *r*^2^ *>* 0.6 were identified as the candidate pruning set, a common threshold for GWAS LD pruning[61]. When two features were found to be in LD, the feature with the lower SHAP z-score was pruned, retaining the feature with greater evidence of predictive importance.

For feature encoding (G*×*G and G+G) pairs, LD was evaluated between and across SNP pairs separately in which pairs (i.e. (*A*_1_*, A*_2_) and (*B*_1_*, B*_2_)) were considered LD-redundant under two cases.*Case 1 = shared SNP* : if the pairs share exactly one SNP (e.g. *A*_1_ = *B*_1_), redundancy was declared if the non-shared partner SNPs were in LD (*A*_2_ *↔ B*_2_); only the partners need be correlated because the shared locus is identical. *Case 2 = no shared SNP* : redundancy required within-pair LD on both sides (*A*_1_ *↔ A*_2_ and *B*_1_ *↔ B*_2_) together with cross-pair LD in either the anti-diagonal pattern (*A*_1_ *↔ B*_2_ and *B*_1_ *↔ A*_2_) or the diagonal pattern (*A*_1_ *↔ B*_1_ and *A*_2_ *↔ B*_2_). In each redundant group the lower-ranked pair (by SHAP z-score) was pruned.

For T2D, of the 1.497 G+G pairs entering this step, 455 G+G were removed; of 13,767 G*×*G, 36 were removed; and of 915 G SNPs, 8 were removed. For CD, of the 191 G SNPs 0 were removed, 1 of the 778 G+G pairs entering this step were removed, and 10 of the 4,747 G*×*G were removed.

#### 4.5.3 G*×*G*×*E and G+G*×*E feature discovery

Pairwise interactions between environmental factors (E) and reduced genetic features and HLA loci were evaluated using gradient-boosted random forests, trained separately for each E. For each model, the training matrix consisted of the *∼*235K training individuals and the reduced G, G+G, G*×*G and HLA feature sets (Methods 4.5.1). Feature importance was assessed using Shapley Z-scores[57, 62], and environmental interaction terms were retained when their Shapley values exceeded those of their constituent features. To further exclude interaction features driven predominantly by environmental effects, we applied a second post-L1-penalized (Lasso) filtering step: interaction terms were retained when their absolute *β* coefficients exceeded individual genetic features in the combined model. Both criteria compare effect magnitudes and do not constitute a formal test of statistical interaction; features passing them are therefore described as environmental interaction *encodings*. The numbers of features retained at each step are reported in the Results and in Extended Tables S2 (T2D) and S14 (CD).

For each clinical measure *k*, an interaction feature was formed for every candidate genetic feature *j* as the elementwise product of the two columns across all training individuals (cases and controls),

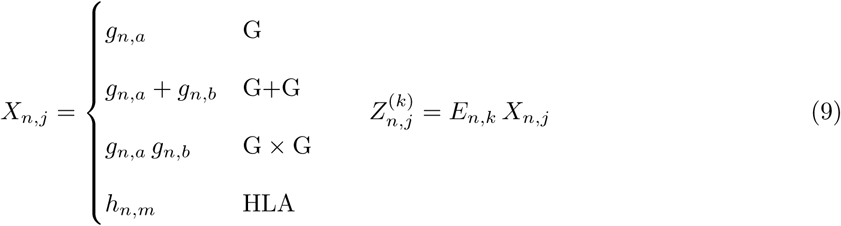

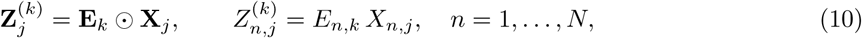

where *⊙* denotes elementwise multiplication. A separate gradient-boosted classifier was then trained for each *k* on a design matrix containing the clinical measure alone, all interaction features, and all genetic features alone,

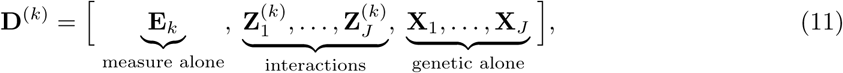

so that each interaction competes directly with both of its constituents. An interaction feature was retained only if its standardised Shapley importance *z* (Methods 4.5.4) exceeded that of both constituents in the same model:

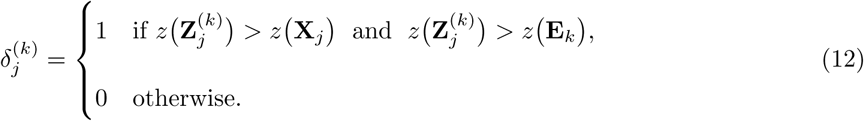

#### 4.5.4 Feature ranking using Shapley importance

Shapley values quantify the contribution of each feature to model performance by averaging marginal contributions across all feature combinations. For all retained G, G+G, G*×*G, and G*×*G*×*E features, Shapley values were calculated using the validation set with fasttreeshap[63] applied to GBH models. Feature z-scores were computed, and features with abs(*z >* 1.99) were retained for final PRS modelling (Table 2).

#### 4.5.5 Confirmatory departure-from-additivity analysis

Shapley importance and L1 selection identify features that contribute to prediction. Neither establishes statistical interaction, because the final models encode each locus pair as a single product or sum term without its constituent main effects. We therefore tested departure from additivity directly, in models that include those main effects.

The analysis was performed separately for each phenotype, on the T2D training cohort (*n* = 235,956; 18,873 cases) and the CD training cohort (*n* = 235,957; 2,622 cases). Each retained pair was re-examined using

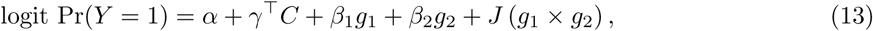

where *g*_1_ and *g*_2_ are the constituent allele counts and *C* comprises age, sex and the first ten principal components. Because both main effects are present, *J* estimates only what the product term explains beyond them: the departure from additivity. Environmental interaction features were tested in the corresponding hierarchy, with *E ×* (*g*_1_ + *g*_2_) requiring a model containing *E*, *g*_1_ and *g*_2_, and *E × g*_1_ *× g*_2_ requiring one that additionally contains *g*_1_*g*_2_, *E × g*_1_ and *E × g*_2_.

Constituents were mean-centered before features were combined. Centering leaves *J* , its standard error and its *p*-value unchanged, since the model is only reparameterised. What it changes is the meaning of the lower-order terms. Without centering, the product is strongly correlated with its constituents (median *|r|* = 0.63) and *β*_1_ estimates the effect of the first locus when the second is absent; after centering the correlation is negligible (median *|r|* = 0.004) and *β*_1_ estimates that effect at the mean of the second locus.

Significance of *J* was assessed by a Wald test and corrected using the Benjamini-Hochberg procedure across the features tested within each model. Pairs for which fewer than 2,000 individuals carried alleles at both loci were not tested, since the product term is identified only on double carriers; this excluded 187 of 1,088 PRS*_GxG_* pairs in T2D and 1,000 of 2,949 in CD, the difference reflecting lower allele frequencies among the CD pairs. This analysis reused the already-selected candidate features and did not repeat the genome-wide pairwise search.

### 4.6 Model Training and ePRS calculations

#### 4.6.1 Association analysis using L1-penalised regression

L1-penalised (Lasso) regression with 5-fold cross-validation was used to reduce overfitting. Models were adjusted for unpenalised covariates obtained from the UK Biobank (age, sex, and the first 10 principal components). In addition to an unpenalised covariate-only baseline, seven models were trained with 181 imputed HLA loci and: (i) linear genetic effects (G), (ii) G+G interaction-informed multilocus additive burden, (iii) G*×*G epistasis-informed product, (iv) combined G+(G+G), (v) combined G+(G*×*G), (vi) G+G burden combined with cardiometabolic environmental interactions (G+G*×*E), and (vii) G*×*G product combined with cardiometabolic environmental interactions (G*×*G*×*E). Statistically redundant models were excluded from downstream analysis. All analyses were implemented in the *glmnet* package in R.

#### 4.6.2 ePRS calculations

Non-zero coefficients from all trained models were used to construct individual-level ePRS calculations according to a generalised additive formula (Equation 14), yielding PRS*_G_*, PRS*_G_*_+*G*_, PRS*_GxG_*, PRS*_G_*_+(*G*+*G*)_, PRS*_G_*_+(*GxG*)_, PRS*_G_*_+*GxE*_, and PRS*_GxGxE_* (Extended Tables S2 and S14 list T2D and CD features, respectively). ePRS distributions were standardized using the StandardScaler function in *scikit-learn*. Model performance was evaluated in the validation set by comparing all models using incremental R^2^, odds ratios, AUC, precision, and recall [64] (Table 2). Statistically distinct models were retained for case identification analysis (4.7).

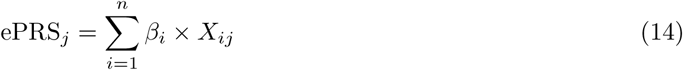

where *j* indexes individuals, *n* is the number of non-zero features, *β_i_* is the L1-regularized coefficient for feature *i*, and *X_ij_* is the feature value for individual *j*: genotype dosage (0, 1, 2) for PRS*_G_*; pairwise allele count sum *X_kj_* + *X_lj_* for PRS_*G*+*G*_; pairwise allele count product *X_kj_* × *X_lj_* for PRS*_GxG_*; combined main and interaction effects for combined models; centered environment times summed allele count (*X_kj_* + *X_lj_*) × *~E_mj_* for PRS*_G+GxE_*; or centered environment times allele product *X_kj_* × *X_lj_* × *~E_mj_* for PRS*_GxGxE_* .

### 4.7 Statistical identification of distinct ePRS models

To avoid redundant reporting of models that capture the same high-risk individuals, all ePRS models were evaluated pairwise and a minimal set of statistically distinct models was retained for downstream analyses. The procedure comprised three stages: pairwise statistical characterisation, a multi-criterion distinctness decision, and a rule-based redundancy resolution.

#### Pairwise statistical characterisation

For each ordered pair of ePRS models, comparisons were performed within cases (affected individuals only) at a fixed high-risk threshold corresponding to the top 20% of each score distribution (the 80th percentile), the same threshold used to define high-risk groups throughout. Specifically, a 2 *×* 2 contingency table was constructed recording the number of cases classified as high-risk by both models, by each model exclusively, and by neither. From this table the following metrics were derived:

i. **McNemar’s test** (asymptotic) to evaluate whether the two models identified significantly different numbers of high-risk cases;
ii. **Cohen’s Kappa (***κ*) to quantify overall agreement in high-risk classification;
iii. the **Jaccard index** *J* = *|A ∩ B| / |A ∪ B|* to measure the fractional overlap of the respective high-risk sets;
iv. a **discordance proportion** with 95% confidence interval calculated with Wilson score interval.

In addition, using the full validation cohort (cases and controls), Pearson and Spearman correlations of the continuous scaled PRS scores were computed within the union of the two models’ high-risk sets, a **paired** *t***-test** was applied to the same subset, and **DeLong’s non-parametric test** was used to compare the areas under the receiver-operating-characteristic (ROC) curves (AUCs) of the two models.

#### Multi-criterion distinctness rule

A pair of models was declared **statistically distinct** if at least two of the following three criteria were simultaneously satisfied:

1. **Low classification agreement:** Cohen’s *κ <* 0.40 (fair or worse agreement);
2. **Low set overlap:** Jaccard index *<* 0.50 (more than half of the combined high-risk set is exclusive to one model);
3. **Different discriminative ability:** DeLong *p <* 0.05 and |ΔAUC*| >* 0.05.

If fewer than two criteria were met, the pair was declared **redundant**.

#### Redundancy resolution

When a pair was declared redundant, one model was removed according to a priority rule. First, a **substring-containment rule** was applied: if one model’s identifier was a proper component of the other’s (e.g., *epi* within *epi +main*). If neither model was a substring of the other, the model with the **lower AUC** was removed as a performance-based fallback. This process was applied iteratively across all pairwise comparisons to yield a minimal, non-redundant set of ePRS models. All downstream visualisations and performance comparisons were restricted to this filtered model set.

### 4.8 Cohort Definition and Important Feature Discovery

Using the holdout set, Individuals were stratified into five ePRS-defined high-risk analytical cohorts. To prevent information leakage, ePRS_Comp_ was excluded from analyses identifying model-specific features.

Three training strata within each cohort were used:

Individuals were assigned to two training strata, each subject to directional quality filters applied prior to enrichment:

- **HighCases**: top-20% T2D cases; only features with OR *>* 1 (risk-elevating direction, *β >* 0) were retained.
- **ExclusiveHighCases**: top-20% T2D cases in exactly one cohort (i.e. n exclusive cohorts = 1) with OR *>* 1; OR was derived as exp(*β*^^^) when not directly available from model output.
- **LowRiskControls**: bottom 20 % of T2D controls in the same PRS distribution.

Important features were identified separately for two data modalities:

#### Genomic features

The holdout set was used to discover cohort-exclusive genomic features derived from pre-computed SHAP (SHapley Additive exPlanations) values obtained from gradient-boosted ensemble classifiers trained to discriminate high-risk cases from controls. A feature was retained if it received a non-zero SHAP contribution in HighCases training partition which included all high-risk cases identified across ePRS models, many of which were identified in more than model, or a smaller subset of high-risk cases exclusively identified in only one model ExclusiveHighCases). The magnitude of each genomic feature’s effect was summarised by its odds ratio (OR), derived as exp(*β*^^^) from the model logistic coefficient. Features were classified as genomic if the feature identifier was a SNP or an HLA allele notation (e.g. HLA-DRB1*401, HLA-B*4002), and as clinical otherwise.

#### Clinical features

Again, using the holdout set, we assessed the importance of 72 clinical variables (Extended Table S12) across ePRS-defined high-risk cohorts. Raw, untransformed features were analysed with *<*5% missingness (and excluded from predictive modelling) for descriptive analysis. Given non-normal distributions, differences were evaluated using the Mann–Whitney *U* test with false discovery rate (FDR) correction applied within each comparison.

#### Enrichment analysis

Clinical values were compared between ExclusiveHighCases and within-cohort bottom-20% controls using a two-sided Mann–Whitney *U* test. Both unbalanced and balanced (bootstrap-resampled, size-matched) designs were used, with median *p*-values from bootstrap replicates retained. Effect size was quantified using the rank-biserial correlation (*r*). Features were retained at FDR-adjusted *p <* 0.05 and *|r| ≥* 0.1, with additional filtering for cross-cohort specificity.

#### Cross-cohort comparison

To characterise broader clinical differences, the same framework was applied across two tiers: (i) **HighRisk**, comparing each top-20% cohort to all others, and (ii) **Exclusive-HighRisk**, comparing exclusive top-20% cases to within-cohort bottom-20% controls. FDR correction was applied per tier, with ExclusiveHighRisk results prioritised where both comparisons were significant to preserve cohort specificity.

#### Cross-cohort specificity tiering

To identify features whose importance was cohort-specific rather than globally elevated, a three-tier specificity filter was applied. Tier 1 (strict): a feature was significant in the target cohort and not significant in any other cohort. Tier 2 (majority): a feature was significant in the target cohort and non-significant in at least 60% of the remaining cohorts. Tier 3 (differential): a feature was significant in the target cohort, which exhibited the highest |effect| across all cohorts, with a minimum advantage of Δ*r ≥* 0.1 over the mean of other cohorts. Each feature was assigned exclusively to the cohort with the highest *|r|* or OR among all tier-qualifying cohorts. Across the five analytical cohorts, between 4 and 333 features per cohort passed these criteria, spanning genomic (SHAP-derived, genomic) and clinical (clinical) data feature types.

### 4.9 Functional Cluster Enrichment

#### Published functional cluster definitions

Functional cluster and clinical phenotype annotations for T2D-associated loci were drawn from four published resources [39–42], spanning 5-16 clusters comprising up to 380 loci (full cluster definitions and sources listed in Extended Table S8). Clinical phenotype associations for each cluster were additionally informed by [46, 47], which characterised relationships between individual loci and quantitative clinical traits (HbA_1c_, glucose, BMI, HOMA-B, HOMA-IR, triglycerides, HDL-cholesterol, and others)

#### Feature-to-cluster assignment

Each important feature identified per cohort was matched to functional clusters using the cluster definitions of Udler *et al.* [41], Kim *et al.* [40], Suzuki *et al.* [39], and Smith *et al.* [42], matched to cohort features by normalised string matching on cluster and feature names (full assignments in Extended Table S8). Genomic features were matched via their nearest candidate gene, clinical features were matched via a Mahajan *et al.* [46] clinical-phenotype bridge table. HLA-region alleles were classified as genomic features and matched by gene prefix.

#### Directional concordance

Each feature-cluster pair was scored for concordance: whether a feature’s observed direction of association agreed with its cluster’s expected phenotypic direction. For clinical features, the observed direction was the sign of the rank-biserial correlation *r* between the feature and case/control status (Extended Table S13); expected directions were derived from published cluster phenotype profiles [39–41, 43]. A score of +1 was assigned when directions agreed, *−*1 when they opposed, and 0 when no expectation was available. Genomic loci were always scored +1, since the pipeline retains only risk-elevating (OR *>* 1) variants. Concordance was not used to filter cluster membership; instead it was carried forward as a signed sum into the subtype-level summaries and figures, so that a feature such as BMI can contribute positively to MOD and MARD while contributing negatively elsewhere.

### 4.10 Assignment of Functional Clusters to T2D Subtypes

Functional clusters were mapped onto the five-subtype taxonomy of Ahlqvist *et al.* [44] (SAID, SIDD, SIRD, MOD, MARD), with LADA treated as part of SAID and MODY as part of SIDD, consistent with the literature. The full cluster-to-subtype mapping, with supporting rationale per cluster, is given in Extended Table S10.

HLA-region features were assigned to the HLA-autoimmune cluster, which maps to SAID; the HLA-DQB1 and HLA-DQA1 alleles additionally have direct published support for this link [45]. Seven clusters lacked direct published subtype evidence and were assigned by literature review: residual glycaemic, HLA-autoimmune, ALP-negative, SHBG/LpA, lipoprotein A, cholesterol, and bilirubin (flagged in Extended Table S10).

Two sets of literature-derived direct evidence supplemented, but did not override, these cluster-level assignments: quantitative clinical-phenotype profiles from Suzuki *et al.*, with subtype association declared at a standardised *z*-score *≥* 0.5 relative to the cross-subtype distribution (27 feature-subtype records), and HLA-DQB1/DQA1 calls from Mansour *et al.* (2 records) (Extended Table S8).

#### Permutation test of differential correspondence

Feature counts per cluster are descriptive: a model may contribute many features to a cluster because it selected more features overall, or because the cluster is large. To test whether the observed concentration exceeds chance, model labels were permuted across features while each feature retained its cluster and subtype annotations, since those annotations are properties of the feature rather than of the model that selected it. Permutation therefore breaks only the model-feature association, preserving both the number of features per model and each feature’s annotation profile.

The test statistic is net concordance: the signed sum of concordance values for a model’s features within a cluster or subtype. A single feature can contribute in opposite directions to different subtypes, as BMI does in being positively associated with MOD and MARD and negatively with others, so the signed sum is the quantity of interest. It is used unnormalised: because the permutation preserves each model’s feature count, every cell is compared against a null generated at that model’s own size, which controls for model size without a denominator. Empirical two-sided *p*-values were computed from 10,000 permutations with a +1 correction and adjusted across all tested cells by the Benjamini–Hochberg procedure. Power is bounded by the number of annotated features per model, so non-significance indicates insufficient evidence rather than absence of correspondence.

### 4.11 Composite Score Construction

Two composite scores were constructed from the statistically distinct ePRS models (five for T2D, three for CD; Section 4.7). The fitted ensemble, ePRS_Comp_, is the composite used throughout the downstream analyses. A rule-based union, ePRS_Rule_, requires no combining weights and is reported only as a comparator, so that the contribution of weighting can be separated from that of the underlying scores.

#### Fitted ensemble (ePRS_Comp_)

Each retained ePRS was z-scored using validation-set means and standard deviations, and ePRS_Comp_ was defined as the linear predictor of a logistic regression of disease status on these scores,

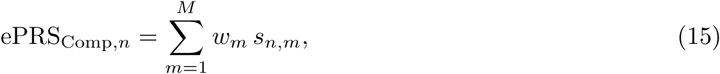

where *s_n,m_* is the standardised score of model *m* for individual *n* and *w_m_* its fitted weight. Weights were estimated in the validation set (Extended Table S3) and applied once to the untouched holdout set, giving a continuous risk estimate that can be thresholded at any operating point. Because most G*×*E scores were tightly clustered around the mean, a small number of upper-tail values became extreme after standardisation. We tested sensitivity to these outliers by capping the highest scores at a fixed percentile, and by replacing each standardised score with its percentile rank across the cohort, which depends only on an individual’s relative position. AUCs under both checks were computed as for the individual ePRS models (Table 2).

#### Rule-based union (ePRS_Rule_)

For each retained model, the 67,402 validation individuals were partitioned into 1,000 quantile bins, and a case rate was computed for each bin relative to overall prevalence. Each of the 33,712 holdout individuals was then assigned a single bin value by three ordered rules: (1) an individual in the top 20% (bin *>* 800) of any model received their maximum bin across models; (2) otherwise, an individual for whom more than 60% of bins were case-enriched (bin case rate *>* prevalence) received their maximum bin; (3) all remaining individuals received their minimum bin. High risk was defined as ePRS_Rule_ *>* 800. Because rule 1 admits anyone flagged by any model, the high-risk group is the union of the model-specific top-20% groups and covers a larger fraction of the cohort than a single model (64.4% in T2D, 45% in CD); rules 2 and 3 affect only the ranking of individuals below this threshold, and therefore the AUC.

#### Comparison

Discrimination of ePRS_Comp_, ePRS_Rule_ and PRS*_G_* was compared with DeLong’s test for correlated ROC curves. Because the two composites flag different proportions of the cohort, case capture was compared at matched coverage: the threshold of ePRS_Comp_ and of PRS*_G_* was lowered until each flagged the same proportion of the cohort as ePRS_Rule_, so that differences reflect ranking quality rather than operating point. Odds ratios compared the top and bottom score deciles.

### 4.12 Evaluation Metrics and Statistical Tests

#### 4.12.1 Statistical evaluation of ePRS performance

Model comparisons were assessed using the DeLong test [37] threshold-free. Odds ratios (ORs) were calculated for the top 10% and 20% of the stratified at-risk population versus the remaining individuals for each ePRS model and each decile versus decile 1 for ePRS_Comp_ and PRS*_G_* in (Figure 5) using contingency tables implemented in statsmodels [65].

Incremental Nagelkerke *R*^2^[66] quantified variance explained by statistically distinct ePRS models, computed as the difference in Nagelkerke *R*^2^ of the trained glm model including the ePRS and covariate versus covariate-only using T2D status as the output only.

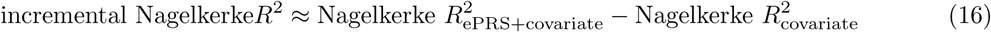

### 4.13 Software and computational environment

Statistical and machine learning analyses were performed using R version 4.4.1[67] (RStudio IDE[68]) and Python version 3.9[69] (scikit-learn[54] for machine learning; statsmodels[65] for statistical analyses).

## Supporting information

ePRS.SupplementaryTable.xls

## Data Availability

All data produced in the present work are contained in the manuscript. All code used in analysis can be found in the GitHub repository: https://github.com/keri/prsInteractive.

## Supplementary Methods: net reclassification

The main text compares ePRS_Comp_ with clinical measures by precision and case yield at a matched classification budget (Figure 6). Net reclassification improvement was additionally computed as a nested comparison, contrasting a clinical model with the same model plus ePRS_Comp_. Both arms were fitted by 5-fold cross-validation and binarised at the reference model’s 80^th^ percentile of predicted risk, so the classification budget is set once by the clinical model and matched in the ePRS_Comp_ model. Against the full clinical model of age, sex, HbA1c, BMI and glucose the NRI was +0.002, and against single measures +0.031 (BMI) and +0.020 (glucose); these are reported in Extended Table S5.

Net reclassification index (NRI) was also used to assess ePRS_Rule_ against each clinical threshold. NRI is the sum of its event and non-event components, the net proportion of cases reclassified upward plus the net proportion of controls reclassified downward. Component standard errors were obtained from the counts reclassified in each direction and combined in quadrature, with 95% confidence intervals given by *±*1.96 standard errors, following Pencina *et al.*[70].

## Data Availability

Results presented here can be found in the Extended Tables 1-15 and available at the UK Biobank under project 86965, “Using Novel Approaches for Improving Polygenic Risk Score Analysis and Filling in Missing Components of Interaction Networks”.

## Code Availability

All code used in analysis can be found in the GitHub repository: https://github.com/keri/prsInteractive.

